# GWAS of Extended Prescription Analgesic Use Identifies Novel Genetic Loci in Chronic Pain

**DOI:** 10.1101/2024.12.02.24318312

**Authors:** Charli E. Harlow, Emeka Uzochukwu, Hazel A. Fernando, Charles E. Mordaunt, Jordan M. Hughey, John D. Eicher, Lara Robinson, Nicholas Bowker, Laurence Howe, Jimmy Liu, Adrian Cortes, Paul Wilson, Usha Gungabissoon, Vicky Tribble, Anthony Nash, Gareth Young, Laura Addis, Chun-Fang Xu, Caleb Webber, Jonathan Davitte, M. Zameel Cader

## Abstract

Pain-related conditions are the leading cause of disability worldwide. Current genome-wide association studies (GWAS) for chronic pain have mainly focused on individual pain-related disorders, which may not optimally capture the phenotype. Here, we applied a novel method of defining chronic pain based on prescription analgesic use (≥90 days) in two large biobanks (UK Biobank and FinnGen). GWAS meta-analyses of 11 prescription-based pain phenotypes identified 140 associations with chronic pain—78 novel (e.g. *ARPP21, CNTNAP2*) and 62 previously reported (e.g. *SLC39A8, DCC, TRPM8*). Integrating these genetic associations with functional data including transcriptome-wide association studies, cell-type and pathway enrichment, and gene enrichment in mouse phenotypes identified novel potential mechanisms involved in chronic pain, implicating oligodendrocyte differentiation, neuronal guidance, endolysosomal function and post-synaptic endosome recycling. Our study showcases how the use of prescription data to identify and characterize pain provides new insights into pain genetics and its underlying biology.

## Introduction

Pain-related conditions are the leading cause of disability worldwide (1). Chronic pain is defined as pain that persists or recurs for 3 months or longer (2). It encompasses a wide range of disorders with complex aetiologies from chronic headache to fibromyalgia. Chronic pain has been reported to affect 13–50% of adults in the UK; and among this group, 10.4–14.3% have moderate-to-severe disabling chronic pain (3). The current range of pharmacological treatments for chronic pain, including opiates, can be associated with harmful effects (4, 5) and/or have limited long-term effectiveness (6). Given the significant unmet medical need for individuals in chronic pain and challenges with existing treatments, there is an urgent need to identify novel targets for chronic pain using human data to allow the development of therapies for improved long-term outcomes.

To date, most genetic studies of pain have been focused on the study of individual pain-related disorders (e.g. migraine, osteoarthritis, back pain). Identified common variants have implicated genes in neurological pathways and inflammation (7). However, focusing analyses on the genetic susceptibility to individual pain-related disorders limits the sample size for studying ‘chronic pain’ in general; and, consequently may miss common biology shared across multiple pain-related diseases. Recent genetic studies leverage ‘pain scores’ based on patient reports or by combining pain traits to increase sample size and the identification of novel pain-associated genetic loci. The ambiguity and heterogeneity in how patients score their pain, however, makes interpretation of these results and their role in the aetiology of pain difficult.

While leveraging diagnosis codes to identify specific pain-related case populations for genetic characterisation is quite straight-forward in its implementation, many individuals never receive a specific pain-related diagnosis and are missed by the diagnosis-focused approach. These individuals may nevertheless seek medical care to repeatedly obtain prescription analgesics to control their chronic pain. Consequently, the availability of prescription data linked to genetic samples in large biobank-data, such as the UK Biobank and FinnGen, provides a unique opportunity for defining chronic pain. In this case, ascertainment is not based on the diagnosis of specific pain-related aetiologies, but rather on the identification of individuals that are seeking treatment for pain relief for extended periods of time. Extended use of analgesics may thus serve as a novel definition of ‘chronic pain’ that: 1) captures both the diagnosed and undiagnosed chronic pain population; 2) increases the total sample size for genetic characterisation of chronic pain; and, 3) aids in the identification of novel pain loci and an improved understanding of underlying biological pathways or mechanisms that are shared across pain-related aetiologies. To our knowledge, no genetic studies to date have yet used analgesic prescriptions to identify and study individuals with chronic pain.

We used a novel operational definition of chronic pain (analgesic use ≥90 days) and conducted genome-wide association studies (GWAS) of pain phenotypes derived from prescription data in individuals of European ancestry in both the UK Biobank (N∼500,000) and FinnGen (N∼500,000). We subsequently completed meta-analyses of the results from these two biobanks to identify common variants associated with these pain phenotypes. We also conducted rare-variant gene-based tests, enabled by the Whole-Genome Sequencing (WGS) data in UK Biobank, to identify rare variants associated with these traits. Additionally, we sought to understand the genetic correlation between our novel definitions of chronic pain with other pain-related aetiologies and psychological disorders (e.g. addiction). Finally, we integrated all our results with genomics analyses from disease-relevant tissues, including gene expression, protein abundance, mouse knockout models, and human disease data, to identify likely effector genes and enhance our understanding of the underlying biological pathways and disease aetiology.

## Results

### Discovery of 140 genetic associations with chronic pain

To increase our understanding of chronic pain, we used prescription data to derive 11 novel chronic pain phenotypes encompassing pain susceptibility, pain severity based on strong opioid use, and pain duration based on time on analgesics using UK Biobank and FinnGen (Figure 1, Supplementary Table 1). We performed GWAS on each of these phenotypes within each dataset and then meta-analysed them together (overall sample sizes: 23,547 – 692,039; Figure 1, Supplementary Table 2). These combined meta-analyses identified 140 associations (*P* < 5 × 10^−8^), of which 78 have not previously been reported as associated with pain phenotypes (Figure 2, Table 1, Table 2). We detected many associations at loci previously reported in the literature (e.g., *SLC39A8, TRPM8)*, demonstrating that our prescription-based definitions of chronic pain are capturing relevant disease information (Table 1) (7–12). Notably, 11 loci previously linked to opioid use disorder and drug metabolism showed association with our prescription defined pain traits (e.g., *SGIP1/PDE4B*, *OPRM1*, *CYP2D6/7*) (13, 14)

**Figure 1:**
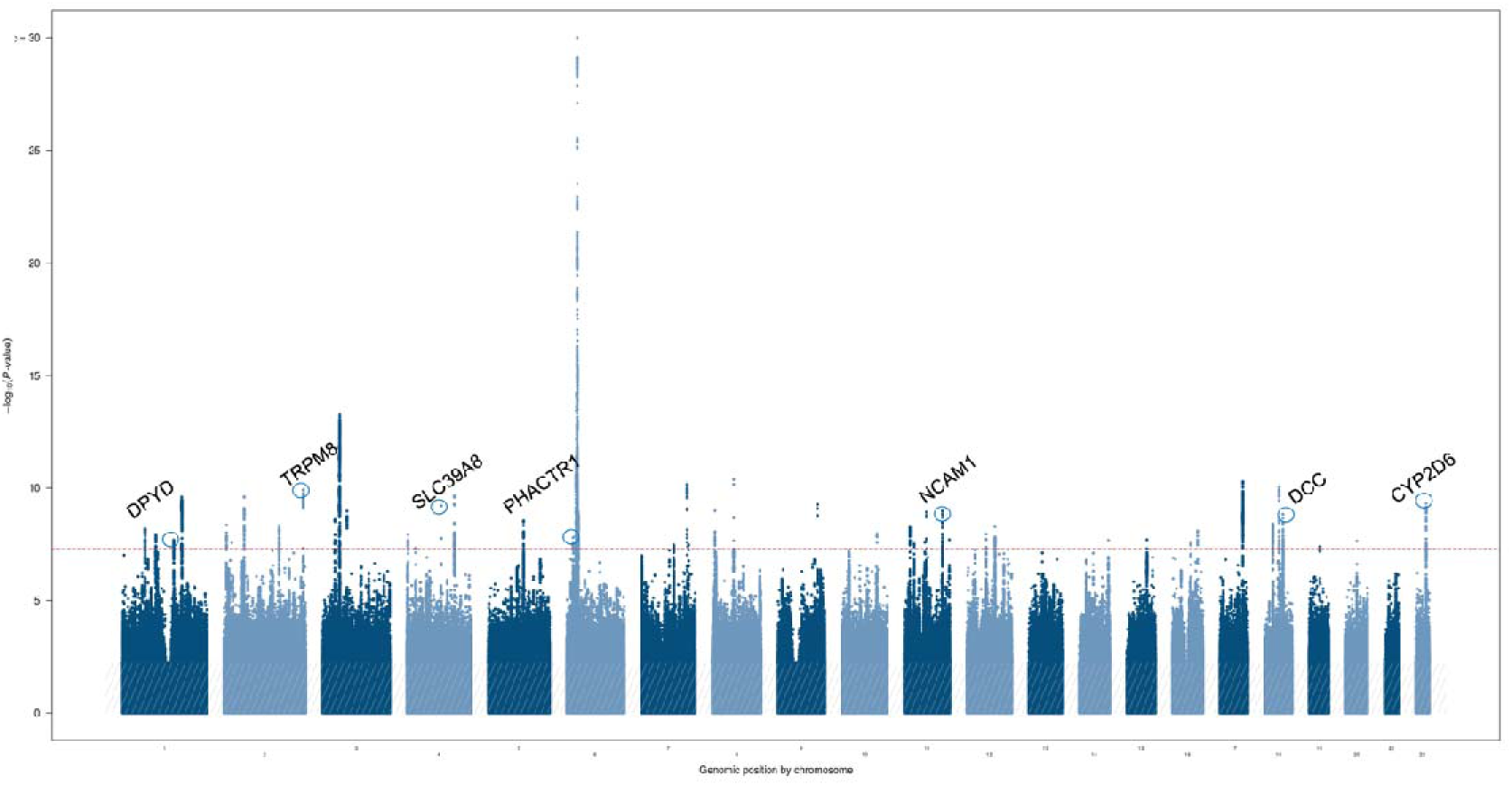
Manhattan plot representing the genetic associations identified through a GWAS meta-analysis of chronic pain, defined using analgesic-use data, in 128,985 cases and 563,054 controls of European descent. Chronic pain was defined using long-term analgesic-use prescription data in UK Bioabnk and Finngen participants. On the x-axis, variants are plotted along the 22 autosomes included in the meta-analysis. The y-axis shows the statistical strength of association from the inverse-variance weighted fixed-effects meta-analysis as the negative log_10_ of the uncorrected P value (P). The horizonal line is the genome-wide significance threshold after correction for multiple testing (P = 5 × 10^−8^). Each dot represents a single nucleotide polymorphism (SNP). Adjacent chromosomes are coloured in different shades of blue. Some well-known pain genes have been annotated by name and circle.

**Figure 2:**
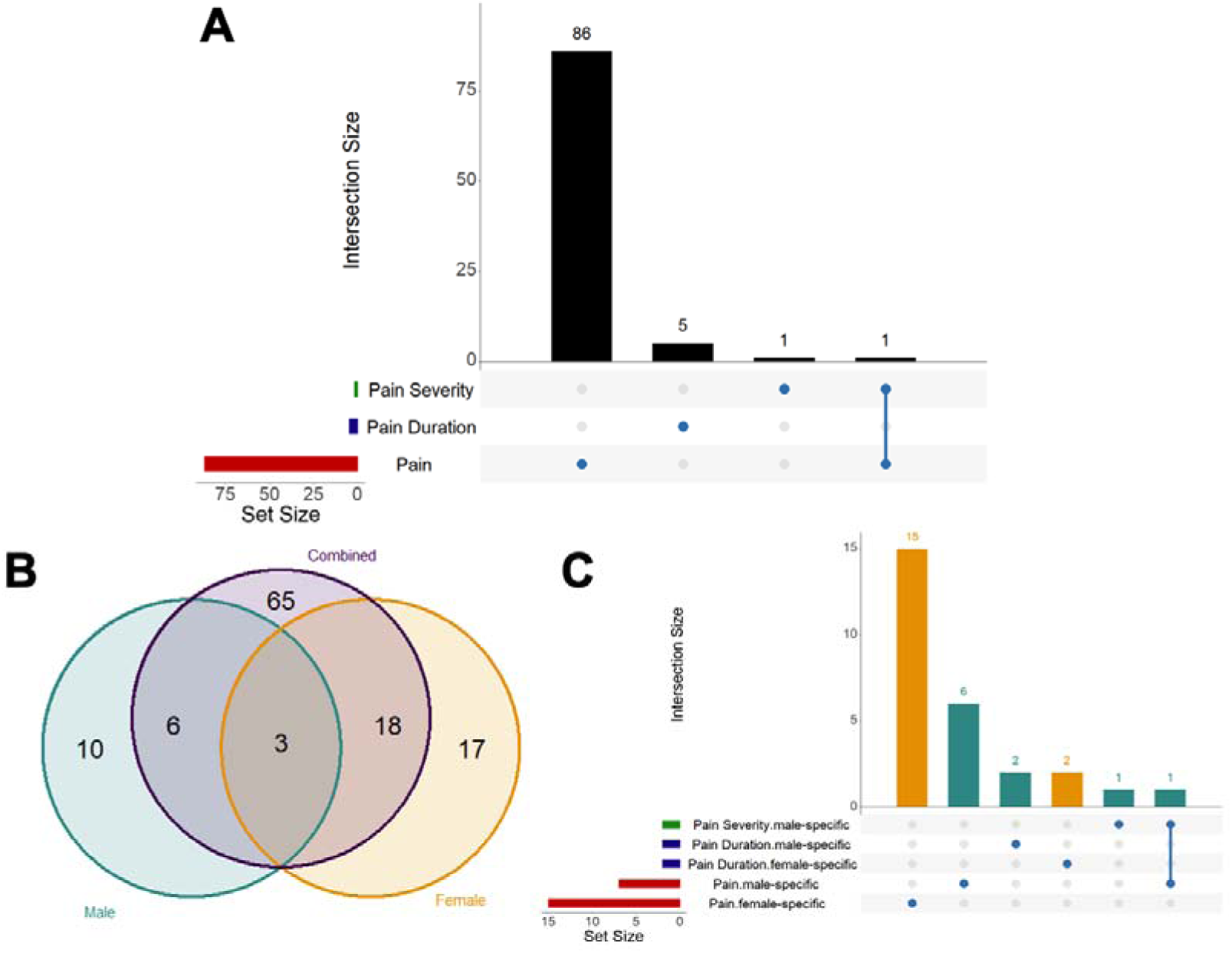
The overlap of genetic loci associated with the three aspects of pain defined using analgesic data - chronic pain susceptibility, pain severity, and pain duration – and in sex-stratified analyses. Overlapping loci were determined based upon an arbitrary 1 mega-base distance between lead variants associated with any of the 11 chronic pain phenotypes. A: Upset plot representing the number of genetic loci associated with chronic pain susceptibility, pain severity, or pain duration. Most loci are associated with chronic pain susceptibility, whilst one locus is specific to pain severity and five genetic loci are specific to pain duration. B: Venn diagram representing the number of genetic loci associated with chronic pain phenotypes in the joint and sex-stratified analyses. There are 10 male-specific loci (representing 11 genome-wide associations) and 17 female-specific loci (representing 21 associations) associated with at least one of our 11 chronic pain phenotypes. C: Upset plot representing the number of male- or female-specific associated with either chronic pain susceptibility, pain severity or pain duration. Most of the male- or female-specific loci, similarly to the joint analyses, are associated with pain susceptibility.

**Table 1:** Summary of the overall genetic findings across each of the chronic pain phenotypes defined using analgesic use prescription data.

| Grouping | Phenotype | Brief Description | Meta-analysis |  | UK Biobank |  | FinnGen |  | Number of independent significant signals | Number of novel signals | Number of previously reported signals |
| --- | --- | --- | --- | --- | --- | --- | --- | --- | --- | --- | --- |
|  |  |  | # Cases | # Controls | # Cases | # Controls | # Cases | # Controls |  |  |  |
| Pain | Chronic pain | Chronic pain patients defined as >90 days prescribed analgesics vs healthy controls (GP linkage and no chronic pain) | 128985 | 563054 | 35550 | 156141 | 93435 | 406913 | 48 | 28 | 20 |
|  | Strong opioid use vs healthy | Strong opioids ever prescribed vs healthy controls (GP linkage and no chronic pain) | 71837 | 563054 | 16505 | 156141 | 55223 | 406913 | 36 | 20 | 16 |
|  | Strong opioid > 90 days vs healthy | Strong opioids prescribed for more than 90 days vs healthy controls (GP linkage and no chronic pain) | 31944 | 563054 | 8100 | 156141 | 23844 | 406913 | 15 | 8 | 7 |
|  | Time on analgesics Q4 vs healthy | Quartile 4 of the time on analgesics compared to participants not receiving analgesics | 32474 | 563054 | 9063 | 156141 | 23411 | 406913 | 21 | 12 | 9 |
|  | Cessation Time Q4 vs healthy | Quartile 4 of the time to stop analgesics compared to participants not receiving analgesics | 32412 | 563054 | 8992 | 156141 | 23420 | 406913 | 13 | 10 | 3 |
| Pain Severity | Strong opioid use > 90 days vs never | Strong opioids ever prescribed vs patients with >90 days prescribed analgesics but never prescribed strong opioids | 31915 | 57085 | 8071 | 18982 | 23844 | 38103 | 2 | 2 | 0 |
|  | Strong opioid >90 days vs <90 days | Strong opioids prescribed for more than 90 days vs Strong opioids prescribed for less than 90 days | 31915 | 23682 | 8071 | 5291 | 23844 | 18391 | 0 | 0 | 0 |
| Pain Duration | Time on analgesics Q4 vs Q1 | Quartile 4 of the time on analgesics compared to Quartile 1 of the time on analgesics | 32432 | 33074 | 9021 | 8709 | 23411 | 24365 | 1 | 1 | 0 |
|  | Time on analgesics Q4 vs Q1-Q3 | Quartile 4 of the time on analgesics compared to all others with time on analgesics | 32432 | 96427 | 9021 | 26403 | 23411 | 70024 | 2 | 2 | 0 |
|  | Cessation Time Q4 vs Q1 | Quartile 4 of the time to stop analgesics compared to Quartile 1 of the time to stop analgesics | 32382 | 32259 | 8962 | 8894 | 23420 | 23365 | 0 | 0 | 0 |
|  | Cessation Time Q4 vs Q1-Q3 | Quartile 4 of the time to stop analgesics compared to all others time to stop analgesics | 32382 | 96477 | 8962 | 26462 | 23420 | 70015 | 2 | 2 | 0 |

**Table 2:**
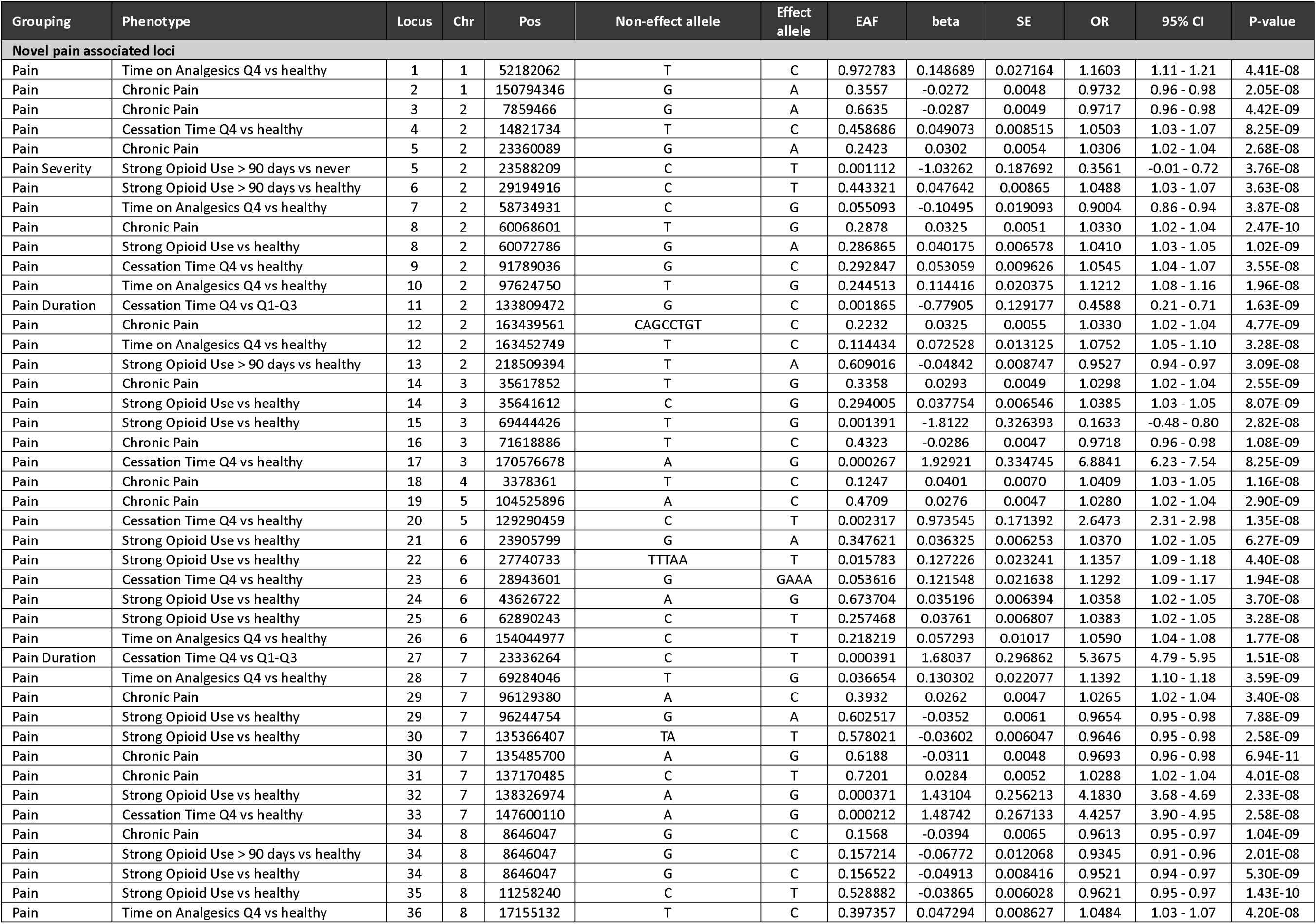
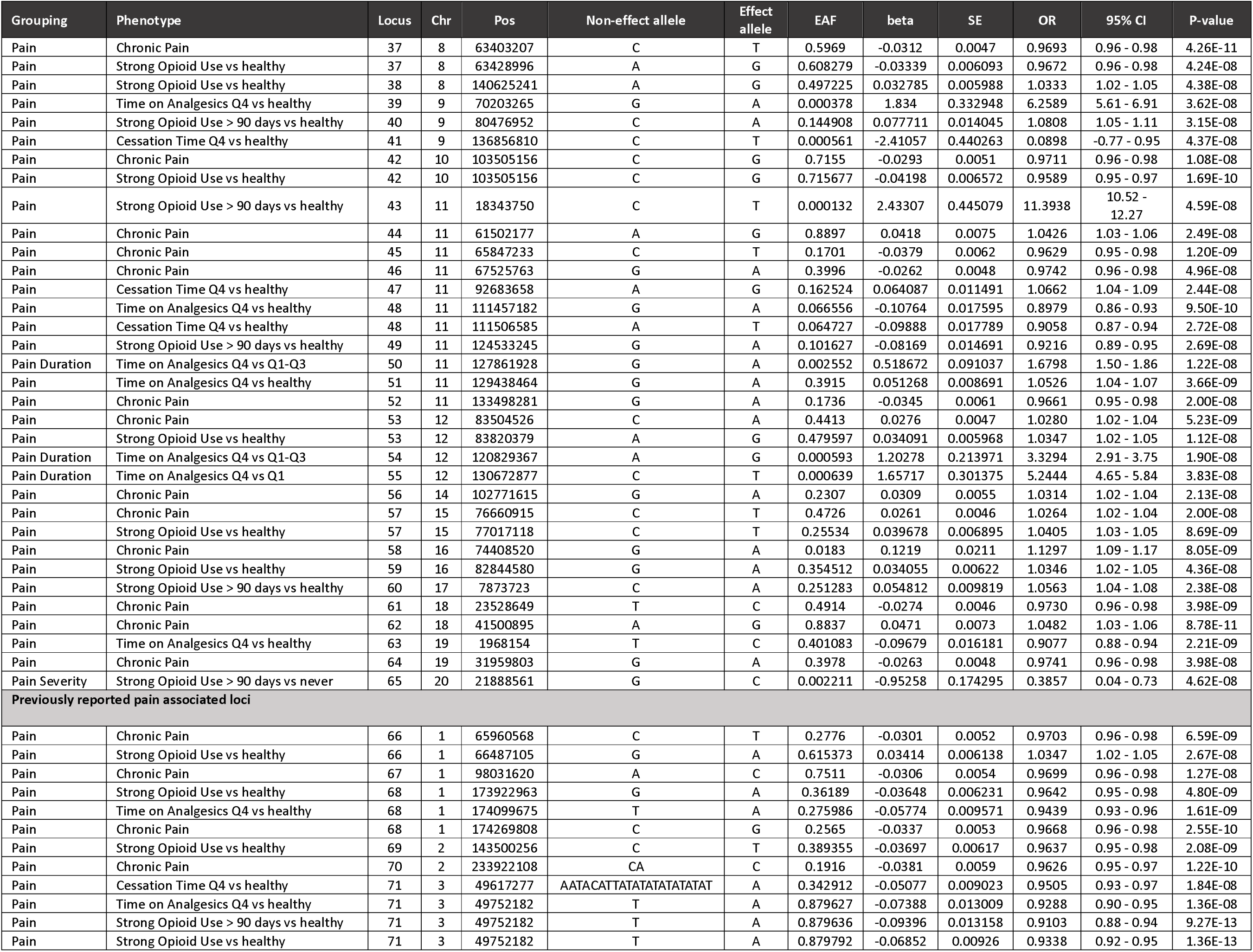
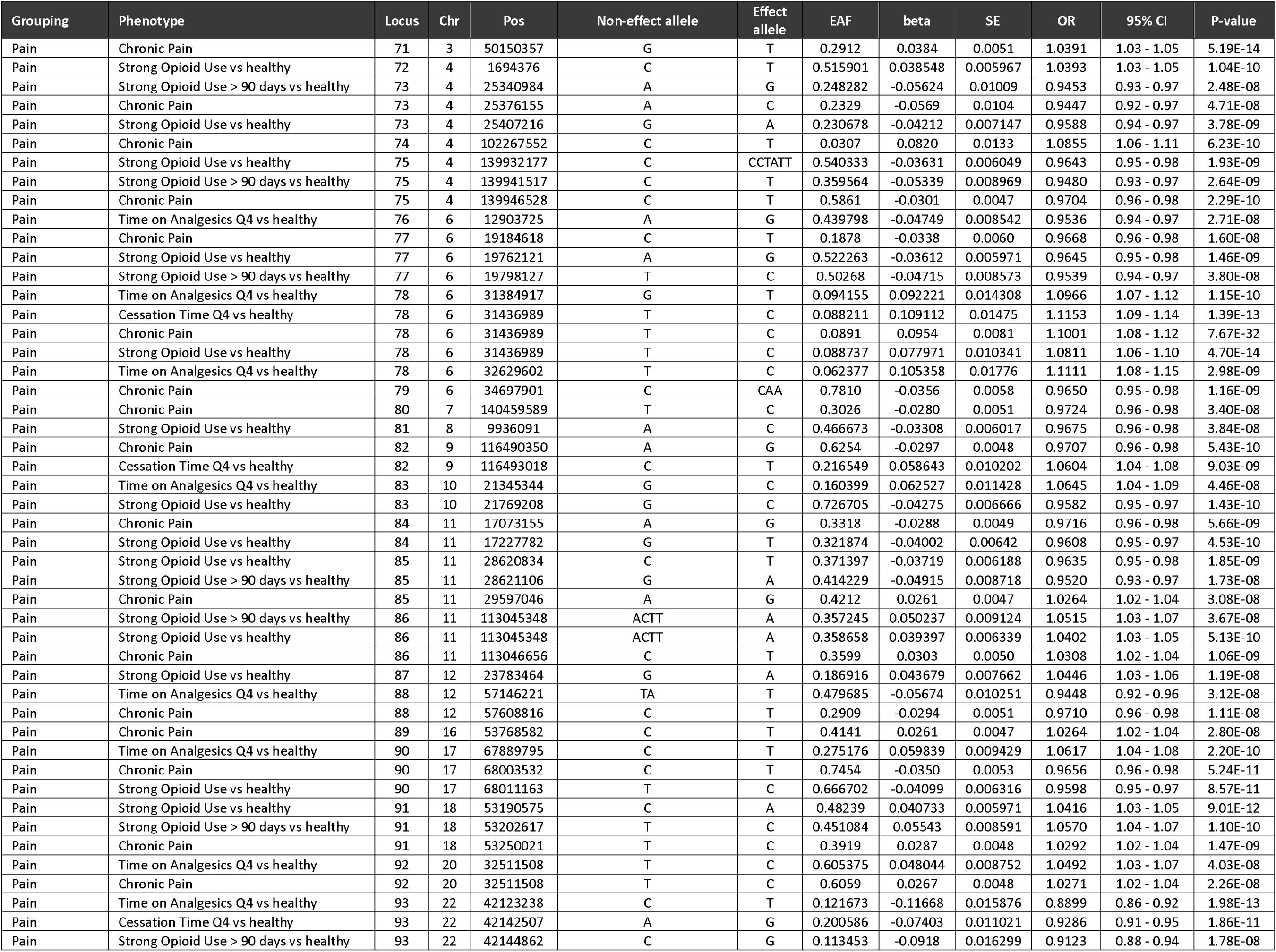
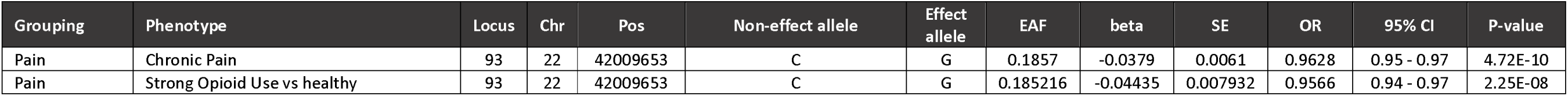
Summary statistics for the 140 independent index variants associated with the chronic pain phenotypes derived using prescription data. Locus number has been defined baased on a 1mb distance from another index variant.

We determined whether the use of prescription data highlighted associations linked to specific pain phenotypes (i.e., susceptibility to chronic pain vs severity vs duration) or were shared across multiple phenotypic domains by assessing the overlap of associations across the three phenotypic definitions. Most loci showed association with only chronic pain susceptibility phenotypes, indicating our phenotype definition is predominantly identifying genes related to central pain mechanisms (Figure 2A, Table 2). When using strong opioid use as a measurement of pain severity, we identified two loci—*PAX1* and *KLHL29* (Figure 2A, Table 2). The *PAX1* locus was specific to only pain severity phenotypes, indicating that it may play a role in pain perception, whilst the *KLHL29* locus was also associated with pain susceptibility highlighting a potential function in both influencing central pain mechanisms and sensitisation. The five genetic loci (*NCKAP5, IGF2BP3, ETS1/KIRREL3, SPPL3, RIMBP2*) associated with pain duration were not associated with pain severity or pain susceptibility implicating their potential roles in the more temporal aspects of pain (Figure 2A, Table 2).

### Sex-stratified analysis identified female- and male-specific GWAS loci for chronic pain

There are noted sex differences in pain perception and treatment with women often experiencing increased pain sensitivity, pain intensity, and recommendation to use opioid treatments (15). We performed sex-stratified GWASs for the 11 derived phenotypes in each cohort, meta-analysed them together, and identified 87 GWAS associations, of which 11 and 21 associations were only seen in males or females, respectively (Figure 2B, Supplementary Table 3, Supplementary Table 4). The slightly larger sample sizes in females compared to males may partially explain why we see a larger number of female-specific loci (Supplementary Table 2). Similar to the analyses not stratified by sex, the majority of the 32 sex-specific loci were associated with chronic pain susceptibility. However, for the pain severity phenotypes, we noted the association of three male-only loci but no female-only loci, suggesting that these genes may play a factor in influencing sex-specific differences in use of strong opioids in pain patients (Figure 2C, Supplementary Table 4).

### Prioritisation of candidate genes for the GWAS associations

To prioritise genes most likely driving the GWAS associations detected in our analyses, we employed a series of complementary computational approaches. First, we annotated the nearest gene based on the distance of the lead genetic variant to the transcription start site or gene-body and found that the majority of GWAS loci mapped to protein-coding genes (Table 3). Next, we determined whether a coding variant with functional impact (i.e., moderate or high impact annotation using the Variant Effect Prediction (VEP) tool) was in the credible set of the GWAS signal. Functional variants impacting 7 known pain genes (*TRPM8, SEMA3F, ANAPC4, SLC39A8, MKRN1, NUCB2, C17orf58, CYP2D6,* and *CYP2D7*) and 4 novel genes (*ALK*, *NPC1, CD6,* and *NAA38)* were identified, implicating that these genes are likely the candidate genes for the GWAS signals (Table 3). Finally, expression and protein quantitative trait loci (eQTL/pQTL) colocalizations were performed to prioritise genes within GWAS loci that may influence phenotypes through modulation of gene and/or protein expression. For 20 loci associated with pain phenotypes, there was evidence of colocalization for eQTL/pQTL for a single gene suggesting that they are likely to be the causal gene within the locus (e.g., *NCAM1, ALK, LRP1)* (Table 3). Prioritisation of the most likely candidate gene was more complex at 37 GWAS loci where there was e/pQTL colocalization for multiple genes within each of the locus (Table 3). We applied a similar prioritisation workflow in the sex-stratified analyses highlighting functional variants impacting 8 genes (*SEMA3F, UHRF1BP1, NUCB2, C17orf58, CYP2D6, CYP2D7*, *PER3*, and *NLRC5*) and evidence of colocalization for only one gene (Supplementary Table 5).

**Table 3:**
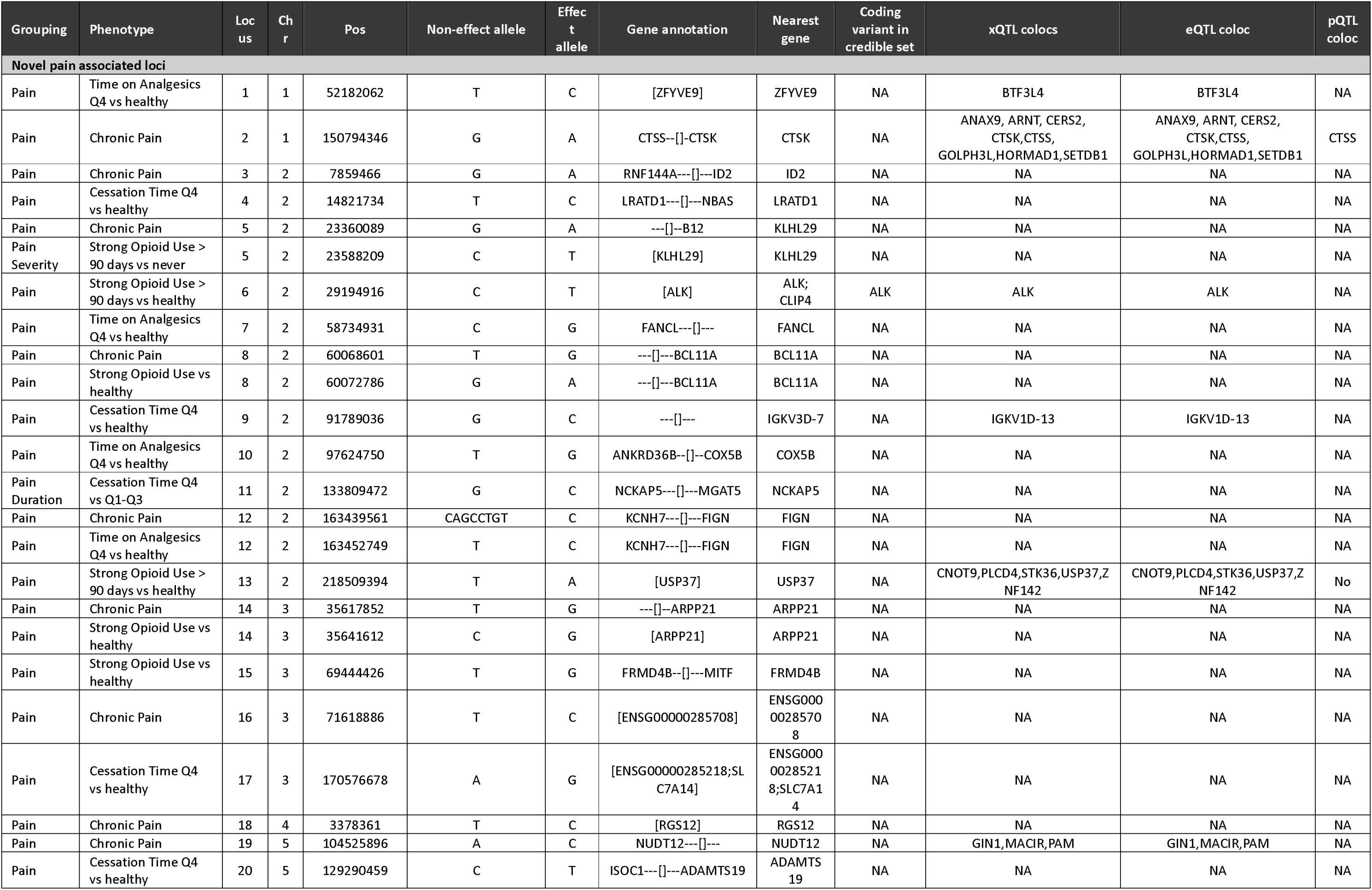
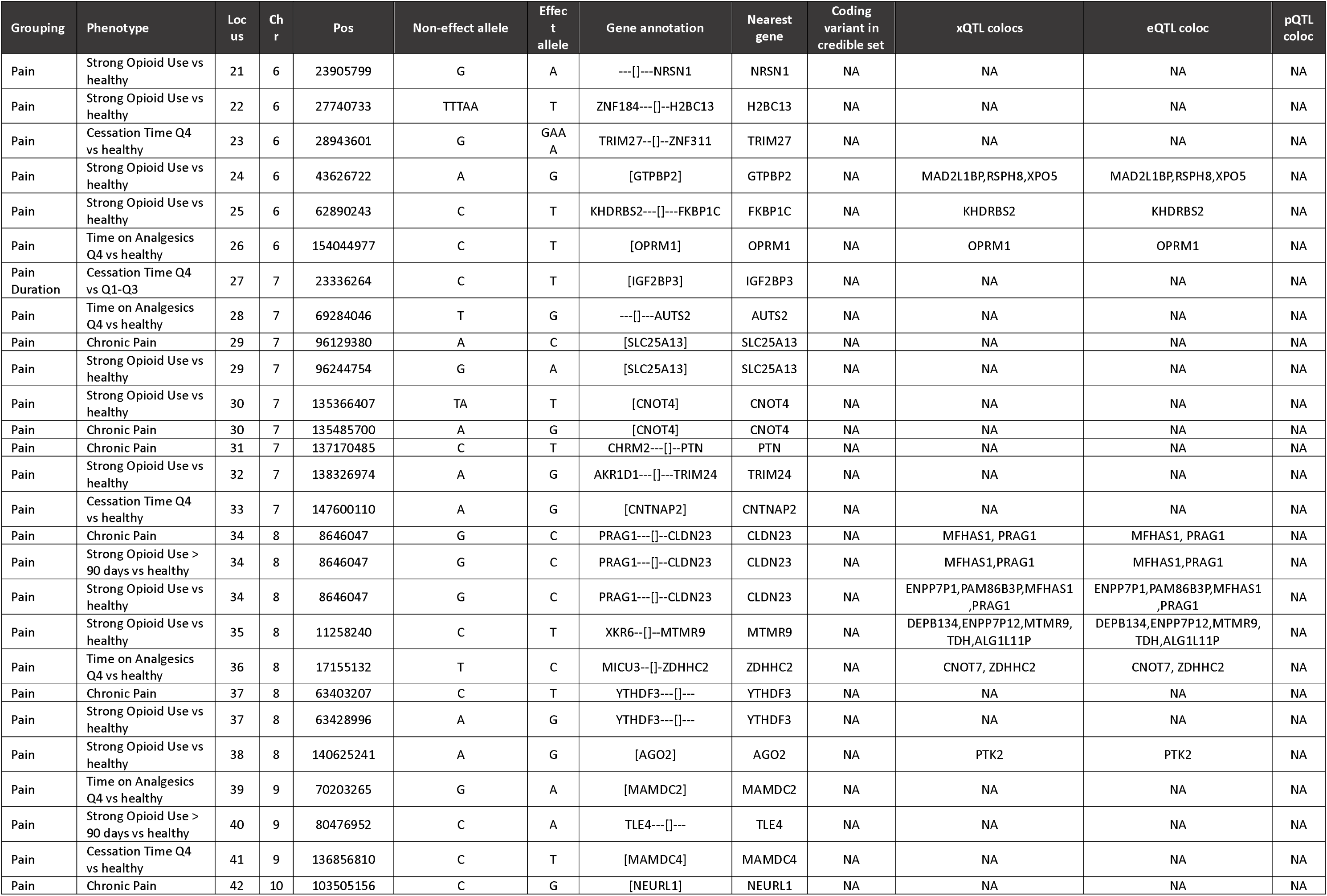
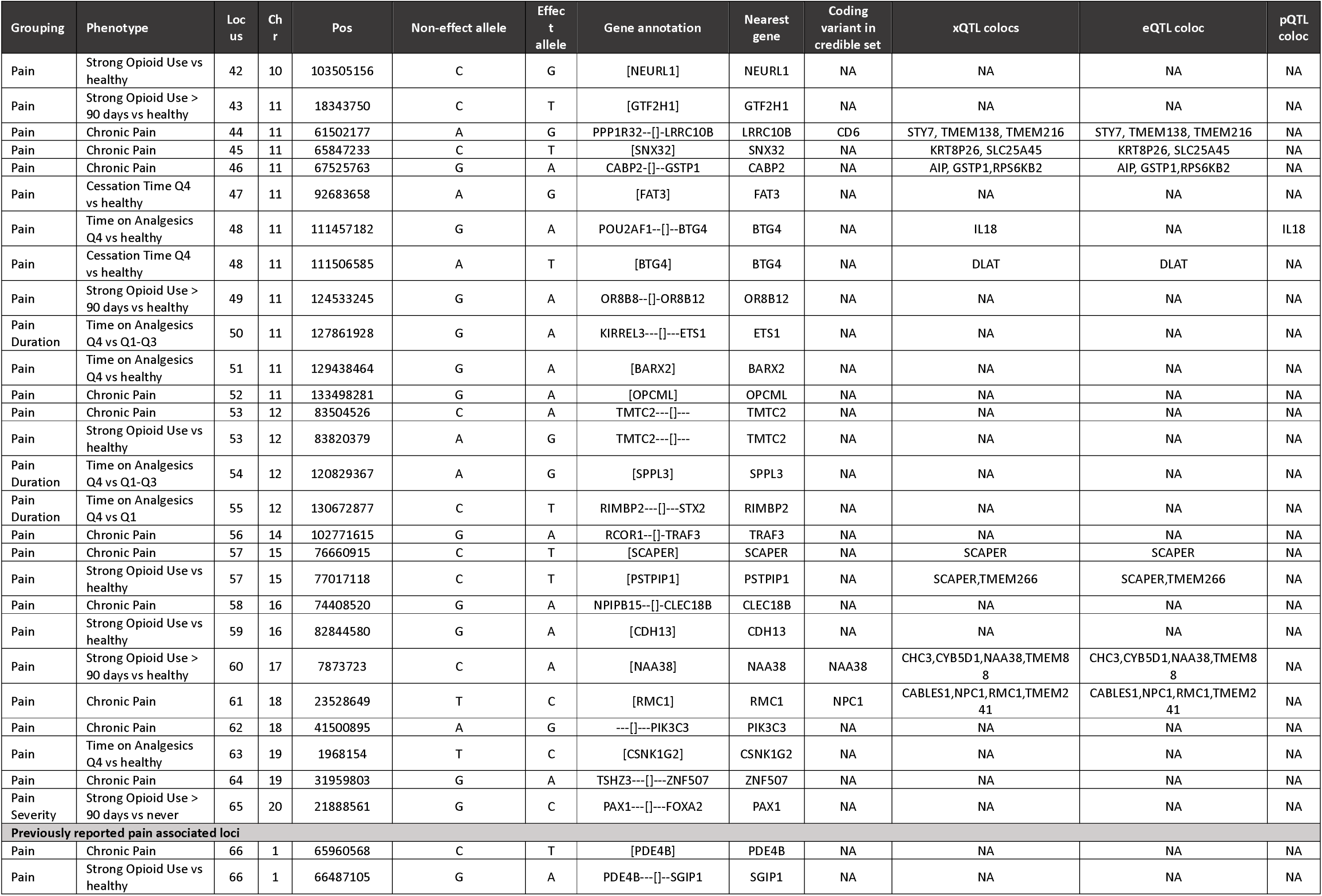
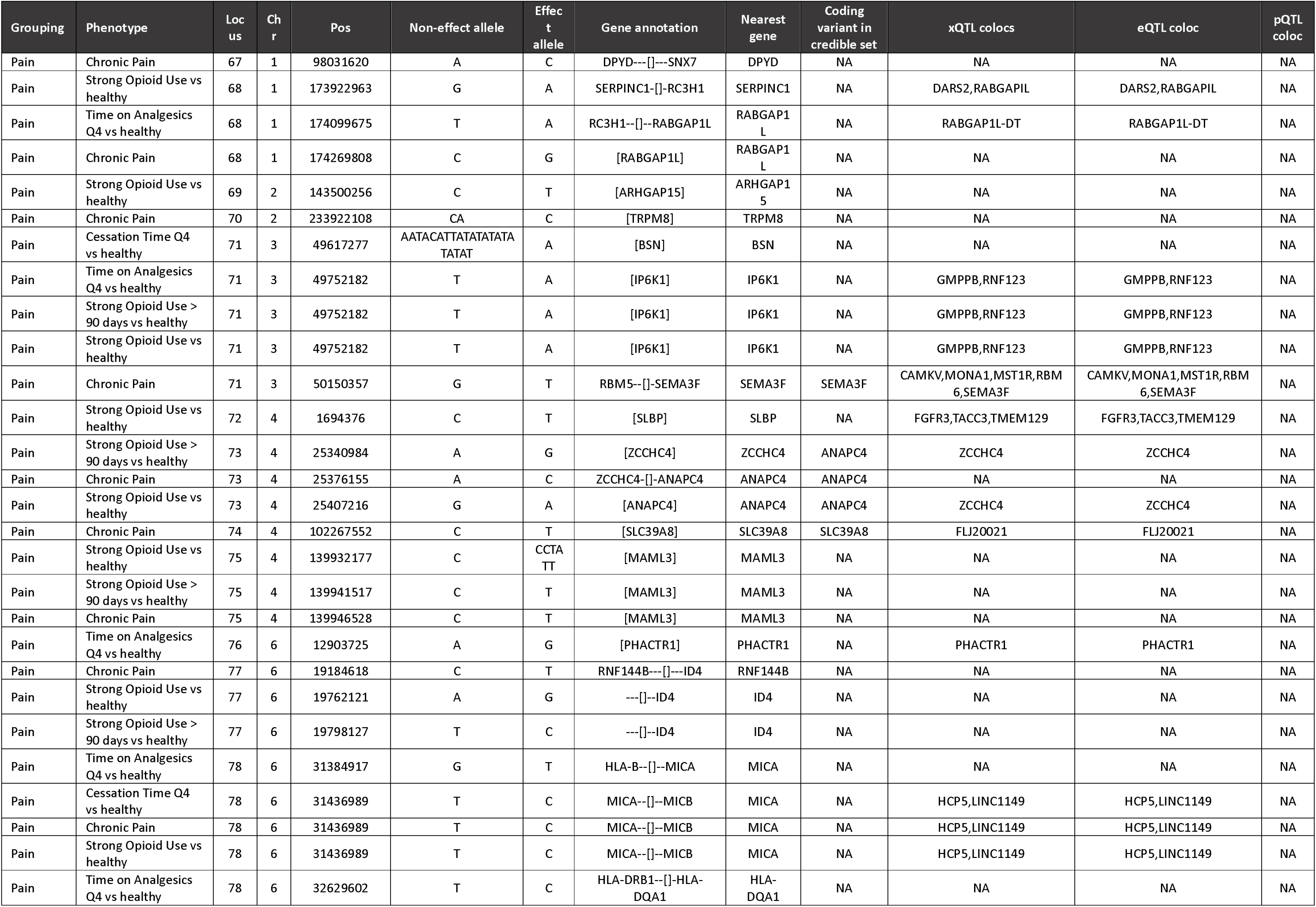
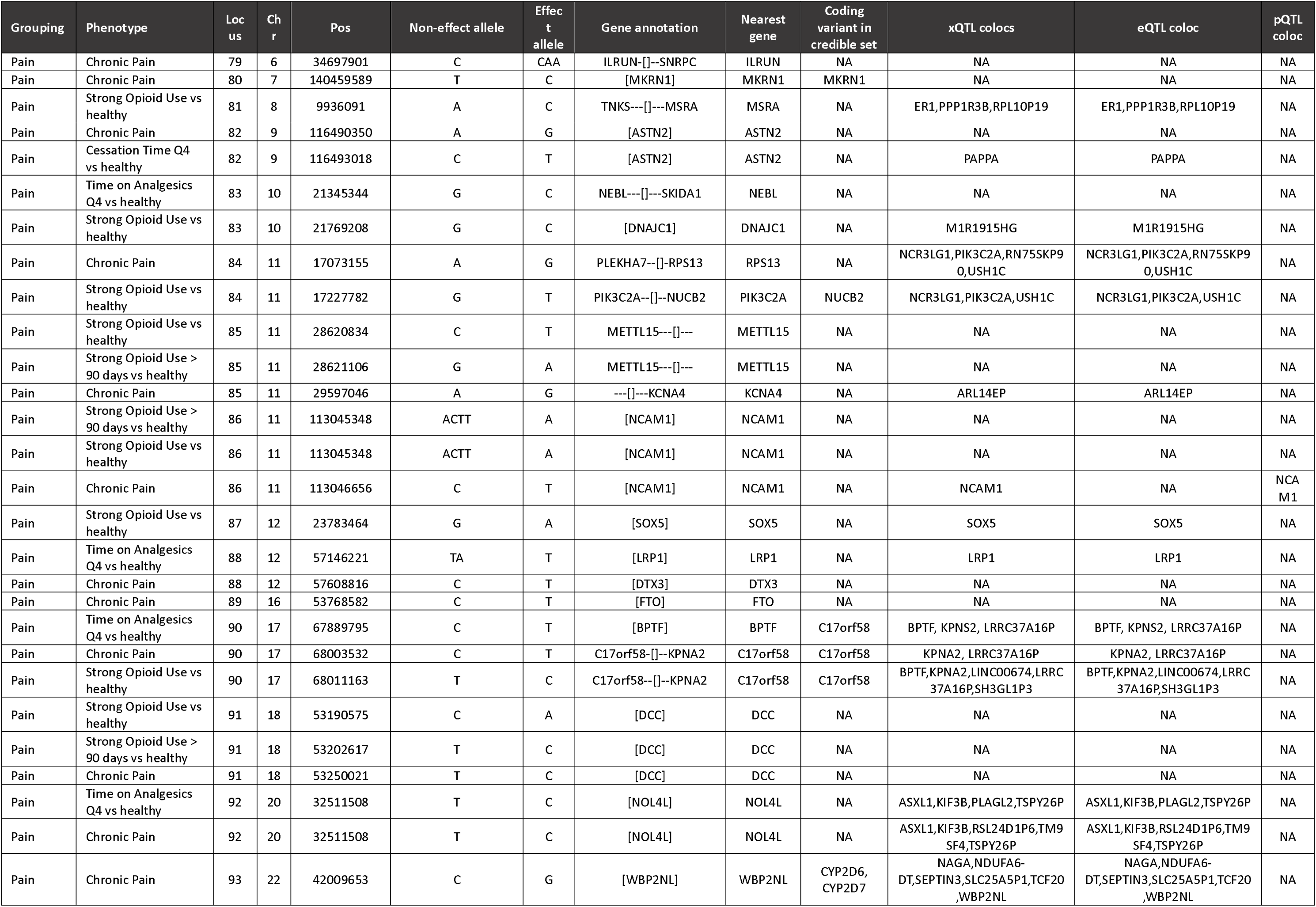
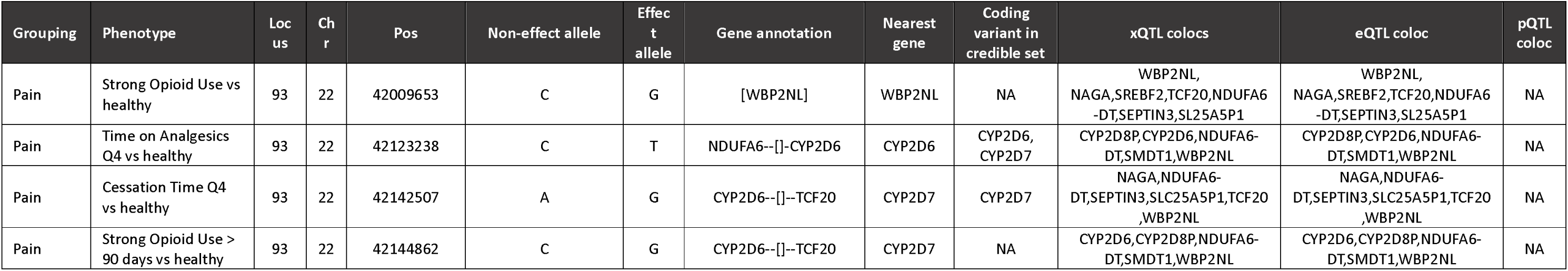
Prioritisation of the most likely causal gene at each of the 140 independent genome-wide significant loci.

### Functional analysis highlights key cell-types and gene-sets linked to chronic pain

We performed a series of functional analyses integrating orthogonal data types (e.g., cell-type specific expression, pathway enrichment, and protein-protein interaction (PPI) networks) to better understand the underlying biology and pathophysiology of pain. We integrated our genetic findings with cell-type gene expression data from cortex, substantia nigra (SN) (16), trigeminal ganglia (TG) (17), dorsal root ganglia (DRG) (18), and gut (18, 19), identifying 12 cell-types associated with chronic pain or migraine (Figure 3A, Supplementary Table 6). The strongest association was seen between cell-type-specific genes from Oligodendrocyte Progenitor Cells (OPCs) in the cortex with pain susceptibility (strong opioid use vs healthy controls: *P* = 7 × 10^−4^) (Figure 3A, Supplementary Table 6, Supplementary Figure 3) with other pain susceptibility associations with neurons of the gut and peptidergic neurons.

**Figure 3:**
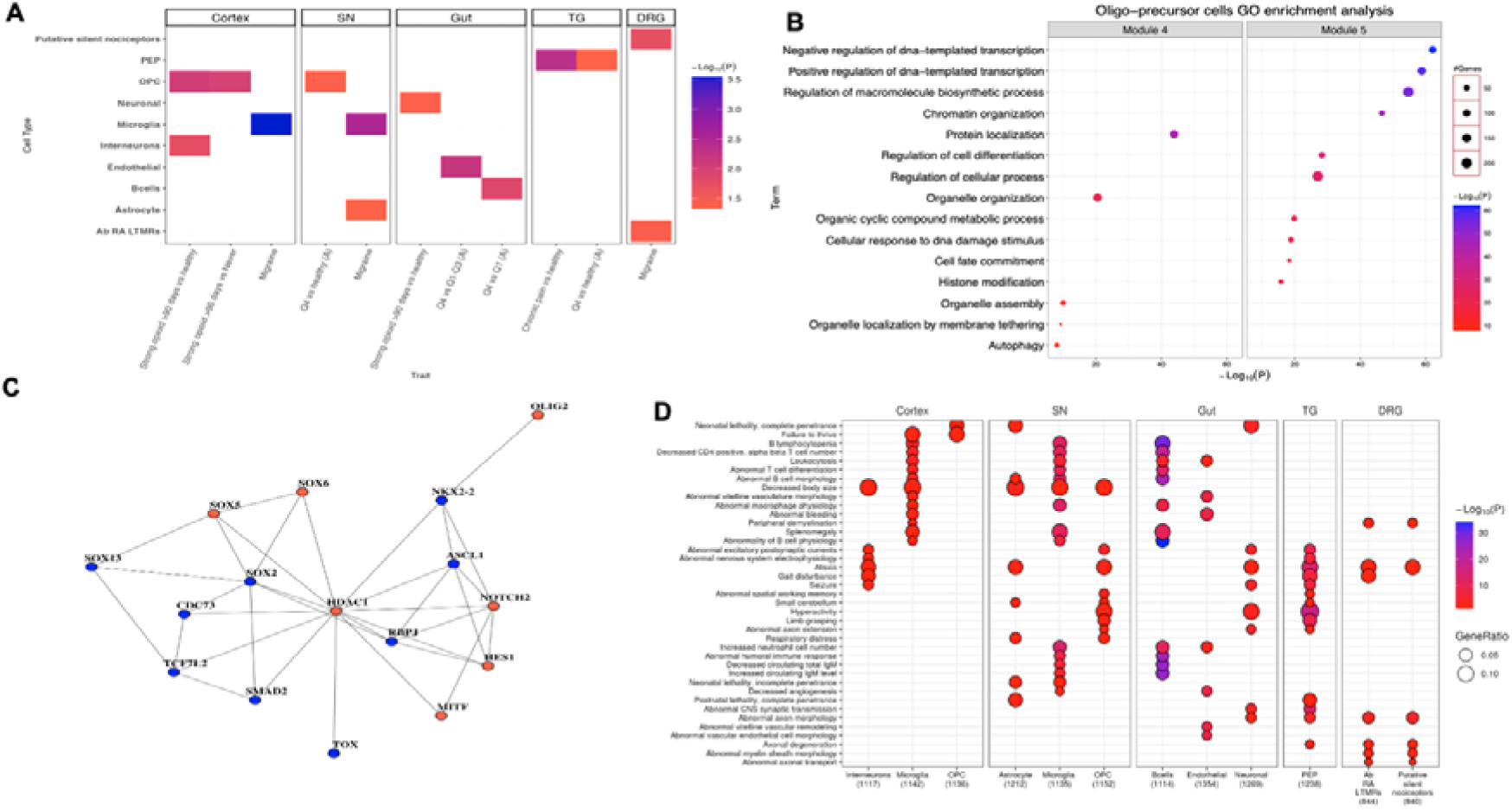
Functional enrichment analysis highlights key cell-types, genes, and pathways implicated in chronic pain. **A)** Cell-types associated with chronic pain phenotypes grouped by central and peripheral nervous systems. Associations are represented as -log10(p-value) with the strength of association shown by the blue – red shading with the blue representing the strongest associations and red the weakest associations. Associations were identified using different methods (MAGMA, LDSC, and AMM) – full details can be seen in Supplementary Figure 3. B) Gene-ontology enriched terms for oligo-precursor cells in the significantly associated modules (Modules 4 & 5). Size of dots represent the number of genes in GO terms in the oligo-precursor cell modules and the colors (red and blue) represent the degree of association. The y-axis are the enriched GO terms. C) Protein-protein interactions (PPI) network for genes present in both cell fate commitment and regulation of cell differentiation. Genes in red are known to be associated with pain disorders from a literature search. D) Mouse phenotype gene sets significantly enriched in chronic pain-associated cell-types. Size of dots represents the proportion of cell-type-specific genes in each mouse phenotype gene-set and the colour represents the degree of association, with blue indicating a stronger association and red a weaker association. The top 5 mouse phenotypes are shown for each cell-type. A square around the circle indicates an additional significant association of the cell-type-specific mouse phenotype genes with one of the chronic pain GWAS. E) PPI network for genes in the abnormal spatial working memory mouse phenotype enriched in GWAS. Genes in red are known to be associated with pain disorders from a literature search. F) PPI network for genes in the abnormal spatial working memory mouse phenotype enriched in GWAS. Genes in red are known to be associated with pain disorders from a literature search.

We next formed PPI modules consisting of the OPC specific expressed genes and re-tested the association between pain phenotypes and OPC specific PPI modules to functionally define the association between pain susceptibility and OPC specific expressed genes. We found that two modules were significanly associated—one functionally annotated as associated with protein localisation (Module 4 *P* = 9 × 10^−22^, Figure 3B) and the other annotated as associated with cell-fate commitment and regulation of cell differentiation (Module 5 *P* = 1.1 × 10^−10^ and *P* = 5.3 × 10^−15^, respectively, Figure 4B). Network analysis of the genes present in cell fate commitment and regulation of cell differentiation highlighted many known genes associated with pain disorders (e.g. *HDAC1, NOTCH2, HES1, SOX5*) as well as novel genes (e.g. *SOX2, SOX13, ASCL1*) (Figure 3C).

**Figure 4:**
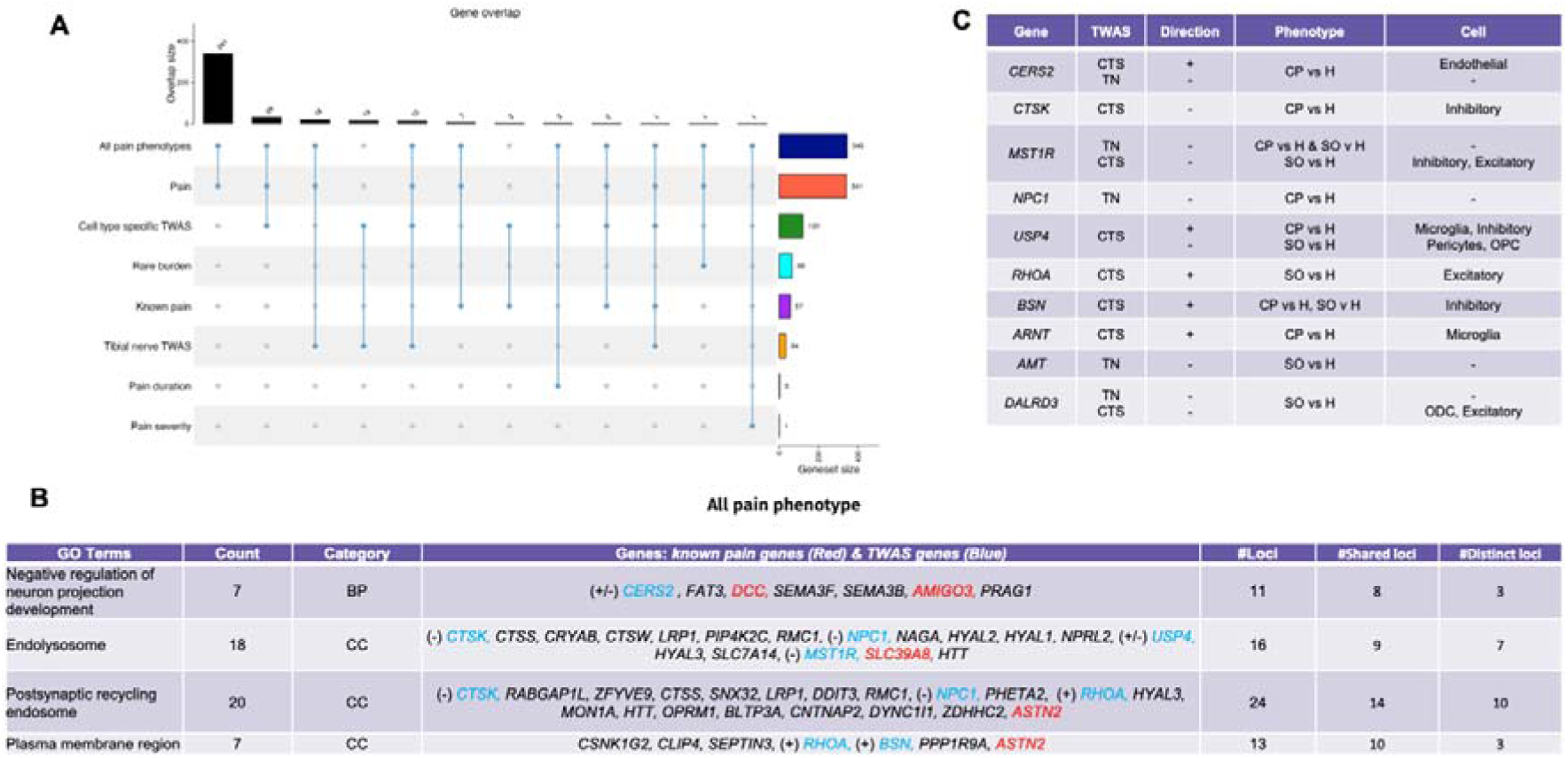
Gene-set overlap and pathway analyses. A) Upset plot representing the overlap between genes identified through our complementary approaches, including GWAS, rare-variant gene-based burden testing, TWAS as well as integrating known pain genes identified by XX et al. and XX et al.. B) The top four most significant GO terms enriched in our gene-set of all pain genes identified through GWAS, gene-based burden testing, and TWAS alongside inclusion of pain genes. Genes identified through TWAS are coloured in blue and directionality inferred for these genes from TWAS is represented by a ‘+’ or ‘-‘ in brackets. ‘+’ indicates that increased gene-expression is associated with increased disease risk whilst ‘-‘ indicates that increased gene expression is associated with decreased disease risk. Known pain genes are coloured Red. C) Summary of the direction of effects inferred from TWAS for some genes identified through TWAS. Abbreviations: CTS = Cell-type-specific, TN = tibial nerve, ODC = Oligodendrocytes, SO vs H = Strong opioid vs healthy, CP vs H = chronic pain vs healthy.

### Functional enrichment analysis with mouse knockout model phenotypes suggests genes, cell-types, and pathways related to chronic pain

To further delineate the underlying pathophysiology of pain and how pain genes may function *in vivo*, we utilised mouse phenotype gene-sets where genetic perturbations are mapped to *in vivo* phenotypes, with a total of 192,000 gene-phenotype pairs (20). We tested for enrichment of these mouse phenotype gene-sets in cell-type-specific genes from 43 cell-types across five tissues (described above—cortex, SN, TG, DRG, and gut). We found significant enrichment for 666 unique mouse phenotype gene-sets (Supplementary Table 7); 403 unique phenotypes are observed in the 12 cell-types we had linked to pain GWASs (Figure 3D). Significantly enriched phenotypes tended to align to the cell-type’s function (e.g., neurological phenotypes associated with neurons and immune phenotypes associated with B cells).

We next tested whether our chronic pain GWASs were enriched for these identified cell-type specific mouse phenotype genes. We found the “strong opioid use” GWAS to be significantly enriched with peptidergic nociceptor-specific genes mapped to bilateral tonic-clonic seizure (*P* = 3.74 × 10^−5^) and abnormal spatial working memory mouse phenotypes (*P* = 1.64 × 10^−6^) (Supplementary Table 8). These two identified gene-sets included glutamate and GABAergic receptors, highlighting a plausible pathway by which these gene sets may potentially contribute to pain mechanisms (Supplementary Table 9, Supplementary Table 10). Other cell-type-specific associations included microglia and gut B cells with analgesic-use duration GWASs, with genes mapped to mouse phenotypes reflecting altered immune cell numbers and function indicating that these pathways may play a role in the temporal aspects of pain (Figure 3D).

### Transcription-Wide Association Study

To determine the genetic relationship between changes in gene expression and chronic pain phenotypes, we performed a Transcriptome-Wide Association Study (TWAS) using eQTLs for eight brain cell-types (21) and the tibial nerve (22) and tested the genetic association of imputed gene expression and our chronic pain phenotypes. This identified 34 tibial nerve and 120 cell-type specific significantly associated genes of which 18 (e.g., *NPC1*, *BPTF*) and 29 (e.g., *BSN, HLA-DRB1*) were also identified through GWAS, respectively. Twelve genes (e.g., *MST1R, GMPPB*) were found in both TWASs as well as the GWAS analyses (Figure 4A, Supplementary Table 11, Supplementary Table 12).

To improve our understanding of the underlying mechanisms and pathways involved in chronic pain, we created a chronic pain gene-set by combining the genes from our chronic pain GWAS, rare variant burden associated genes, and TWAS, alongside 57 previously identified (‘known’) pain genes (7, 23) and performed Gene-Ontology (GO) analysis on this gene-set (Supplementary Table 12). GO analysis highlighted regulation of neuronal projection, endosomal pathways including postsynaptic recycling, and the plasma membrane (Figure 4B). Integration of the TWAS data with the GO findings can help in assigning directionality of pathway changes by providing information on gene expression changes within cell-types. Our results show that direction of effects can be cell-type specific--for example, *CERS2* was significantly associated in endothelial cells and the tibial nerve TWASs with observed effect in oppositive directions (Figure 4C).

### Shared genetics between analgesic-use defined pain phenotypes and other pain-related diseases

We performed LD Score Regression (LDSC) analyses to investigate the degree of genetic heritability and shared genetic architecture between the 11 prescription - based pain phenotypes and 12 GWASs of conventional pain-related traits (fibromyalgia, migraine, and osteoarthritis). This analysis would shed light to the new approach we applied to define pain phenotypes using prescription-based data to identify genes and pathways involved in pain. The sample sizes varied by condition, ranging from 85,708 for fibromyalgia to 1,174,790 for osteoarthritis (Supplementary Table 13). The proportion of variation in chronic pain explained by common genetic variants was similar across the 11 prescription based pain phenotypes (0.002-0.04) (Supplementary Table 14) and to that estimated for migraine, fibromyalgia, and osteoarthritis (0.008 – 0.01) (Supplementary Table 13). The strongest positive genetic correlations were between fibromyalgia and our measures of chronic pain susceptibility, and pain severity (r_g_ ranging from 0.8 to 0.9, *P* < 1 × 10^−5^), whilst no significant genetic correlations were seen between fibromyalgia and pain duration phenotypes (*P* < 1 × 10^−5^) implying that fibromyalgia is more closely related to pain sensitisation. Additionally, moderate positive genetic correlations (r_g_ > |0.5|, *P* < 1 × 10^−5^) were observed between our definition of chronic pain and osteoarthritis and migraine (r_g_ 0.50 - 0.78). To delineate whether these correlations were likely linked to sensitisation or temporal aspects of pain, we compared the level of genetic correlation for pain duration to pain severity phenotypes and found moderate genetic correlations between pain duration phenotypes and migraine only (r_g_ ranging from 0.51 to 0.55) (Figure 5, Supplementary Table 15). For sex-specific genetic effects, stronger genetic correlations tended to be with fibromyalgia in males as opposed to females (Supplementary Figure 4, Supplementary Table 15). Consistent strength of genetic correlations was seen between males and females for pain phenotypes with migraine and osteoarthritis (Supplementary Figure 4, Supplementary Table 15). Similar to the combined analyses, migraine was genetically correlated with pain duration as opposed to pain severity in sex-stratified analyses (Supplementary Figure 4, Supplementary Table 15).

**Figure 5:**
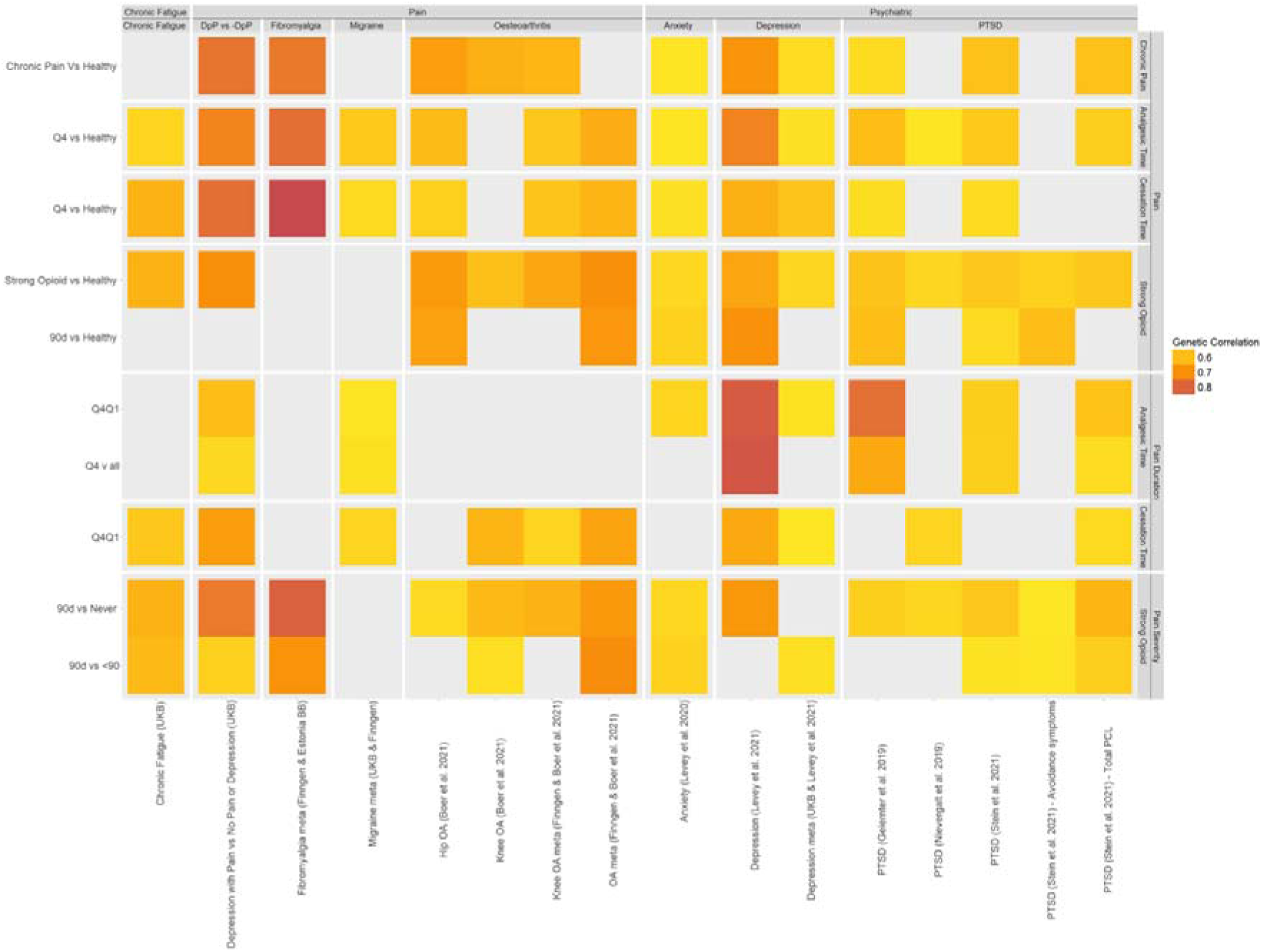
Heat map of the shared genetic effects between chronic pain phenotypes and other pain-related, immune-related and psychiatric conditions. Heat map shows only those results passing a P < 1 × 10^−5^ and with a genetic correlation score > |0.5|. The y-axis shows the chronic pain phenotypes defined using the prescription data clustered by pain susceptibility, pain duration and pain severity. The x-axis shows the phenotypes for which we were assessing genetic correlation clustered by pain, and psychiatric. The darker the orange shading the stronger the genetic correlations. There were no significant genetic correlations between chronic pain phenotypes and immune or anthropometric traits. Abbreviations: DpP = depression with chronic pain, -DpP = no depression or chronic pain

### Shared genetic architecture of pain with neuropsychiatric and immunological traits

Many studies have demonstrated a link between chronic pain and psychiatric conditions, mental health conditions and immune related disorders (24–27). To determine if our chronic pain phenotypes share genetic components with these conditions, we performed LDSC on 21 traits spanning across psychiatric (e.g., bipolar, depression and schizophrenia), immune-related traits (e.g., Crohn’s, Inflammatory Bowel Disease (IBD) and rheumatoid arthritis), and anthropometric traits (e.g., height & weight) (Supplementary Table 14). We selected these GWAS analyses due to their known comorbidity with pain related conditions, the availability of robust GWAS data, and their representation of diverse biological mechanisms. The strongest genetic correlations were found between pain duration phenotypes and depression or post-traumatic stress disorder (PTSD) (r_g_ > |0.8|, *P* < 1 × 10^−5^; Figure 5, Supplementary Table 15). Modest but consistent genetic correlations were also observed across the other pain categories (chronic pain, pain severity) with psychiatric conditions (e.g., anxiety and, PTSD) (Figure 5, Supplementary Table 15). Despite other studies showing a link between chronic pain and immune-related disorders or anthropometric traits implying some shared mechanisms (24–27), we did not find any evidence of significant genetic correlation between chronic pain and these phenotypes (Figure 5, Supplementary Table 15). Chronic fatigue, a multifaceted disorder, was also included in LDSC analyses to determine the degree of genetic overlap with pain as the exact causes of chronic fatigue remain unknown (28). Consistent moderate genetic correlations were observed between chronic fatigue and our three pain phenotypes indicating that some shared genetic factors could influence pain symptoms and treatments given to chronic fatigue patients (Figure 5, Supplementary Table 15).

### Association of rare variants with chronic pain phenotypes

In addition to interrogating common variants associated with chronic pain phenotypes, we performed gene-based association of rare variants testing with chronic pain in UK Biobank. While we did not observe any exome-wide significant associations (*P* < 1 × 10^−6^) with any of our pain traits in UK Biobank, we did identify 66 genes with a suggestive association with chronic pain phenotypes (*P* < 1 × 10^−4^) (Supplementary Table 16). One of the rare variant gene-based suggestive associations (*PIK3C2A*) was also observed in our common variant GWAS of pain susceptibility (*P* = 4.5 × 10^−10^) (Table 3, Supplementary Table 12). The variant-to-gene mapping at this locus was complex with QTL colocalization evidence for multiple genes. However, this gene-based suggestiveassociation between rare LoF variants and risk of chronic pain suggesting that *PIK3C2A* is likely the candidate gene for the association within the locus. We did not observe any rare variant gene-based suggestive associations (*P* < 1 × 10^−4^) with pain severity or pain duration phenotypes (Supplementary Table 16).

## Discussion

We conducted the first genome-wide association meta-analysis of chronic pain defined by analgesic prescription data in up to 1,000,000 individuals of European descent. We identified 140 GWAS associations for chronic pain phenotypes, of which 78 are novel. The number of identified loci is comparable to the number (125) identified by Toikumo *et al.* (11), who focused on pain intensity, and is considerably higher than the 99 loci identified in prior chronic pain GWAS’ (8, 10, 29–32). This new method of defining chronic pain patients via their medication usage allows better characterisation of an otherwise poorly diagnosed and heterogenous phenotype that is not often well-captured by diagnosis-code based approaches, enabling the identification of novel pain association signals (e.g. *CNOT4, CNTNAP2*, *ARRP21*). Most GWAS loci, including all previously reported, were associated with susceptibility to chronic pain rather than severity of pain or duration of analgesic use. Heritability across the pain phenotypes were similar suggesting that the fewer loci found for severity and duration may be due to a lack of statistical power compared to the larger susceptibility definitions.

The chronic pain locus we identified on chromosome 3 p21.31 contains many plausible causal genes (Supplementary Figure 5). Although there is eQTL colocalization evidence for both *RBM6* and *CAMKV* (Table 3), the strongest evidence points to *SEMA3F* for which there are coding variant evidence (Leu503Met, rs1046956:T:A, EAF = 0.72, OR = 0.96, *P* = 1.4 × 10^−13^) and eQTL colocalization evidence. eQTL colocalization suggests that increased *SEMA3F* gene expression associated with increased chronic pain risk. SEMA3F is a class 3 semaphorin which are secreted ligands regulating axon guidance and ensuring appropriate pathfinding of sensory afferents in the spinal cord (33). Semaphorins bind to neuropilins and plexins, and there is growing evidence for the involvement of axon guidance molecules more generally in chronic pain disorders. For example, *DCC,* which encodes the netrin-1 receptor and regulates axon guidance, is repeatedly identified in genetic studies of pain, including our own (34). The neuropilin-1 gene has been identified as a migraine susceptibility gene (35). In mice, Plexin-B1-RhoA signalling (with RhoA also identified in our TWAS - Supplementary Table 11) is implicated in inflammatory pain (36), and in humans an auto-antibody against Plexin-D1 has been proposed as pathogenic in neuropathic pain (37). Furthermore, these candidate genes from our GWAS and TWAS were significantly functionally associated with a common neuronal influence (GO term: Negative Regulation of Neuronal Projection Development, *P* = 7.5 × 10^−4^). Given the strong analgesic effects of therapies targeting the neurotrophic factor, NGF, which nevertheless has been limited by degenerative effects on the joint (38), the evaluation of safer therapeutic opportunities in axonal guidance molecules deserves further exploration.

Sex-stratified analyses revealed 21 female-specific associations and 11 male-specific associations indicating potential sex-specific effects. Similarly to the combined analyses, the majority were associated with susceptibility to chronic pain rather than severity of pain or duration of analgesic use. Sex-stratified loci were only associated with pain severity in males implicating that male pain perception may be more genetically driven than female pain perception.

We found a robust association for *PER3* (Asp852His, rs145213510:G:C, OR = 2.02, P=3.7 × 10^−8^. EAF = 0.002) as a female-specific locus associated with chronic pain susceptibility (Supplementary Table 6, Supplementary Figure 6). *PER3* is a clock gene, which has not previously been associated with pain, but is a risk gene for morning chronotype and insomnia (39, 40); structural polymorphisms are associated with delayed sleep-phase syndrome (41) and mouse *PER3* knock-outs have shorter circadian periods (42). Both human pain sensory thresholds (43) and chronic pain disorders (44) have strong diurnal patterns which may reflect direct involvement of circadian clock genes (45) or the influence of circadian-dependent processes such as sleep-wake cycles (46) on pain mechanisms. Sex-specific effects on circadian rhythm have been reported with women having shorter circadian periods (47). *PER3* may therefore affect pain susceptibility in women indirectly through changing the circadian period or affecting sleep (48) and improvements in circadian alignment through lifestyle changes or melatonin treatments could reduce susceptibility to chronic pain disorders. Further studies are required to determine how the *PER3* missense variant (Asp852His) affects protein function and confirm the likely role *PER3* has in pain.

Additionally, we also identified *OPRM1* as a male-specific locus associated with chronic pain susceptibility and duration with evidence of eQTL colocalization indicating higher OPRM1 mRNA expression is associated with reduced time on analgesics (Supplementary Table 6, Supplementary Figure 7). *OPRM1* encodes the mu-opioid receptor, which is the primary site of action of endogenous opioid peptides (49). We previously showed using transcriptomic data from mouse models of pain, that suppression of the opioid signalling network including Oprm1, is evident in the transition from acute to chronic pain (50). Furthermore, polymorphisms in *OPRM1* are associated with differential response to opioids (51). The male-specific association in our study is interesting and aligns with previous studies showing differential efficacy of opioids in males and females (52, 53) . Genetic risk factors altering the efficacy of opioids could in turn lead to altered use behaviour and dependence. Better understanding of the role of *OPRM1* could improve chronic pain outcomes including developing approaches to reduce the risk of prolonged use of exogenous opioids.

Gene burden-test analysis, despite not identifying any statistically significant associations (*P* < 1 × 10^− 6^), did identify 66 genes suggestively associated with chronic pain (at *P* < 1 × 10^−4^). This is the largest such study to date and identifies genes not previously reported as potentially associated with chronic pain. The overlap with loci identified through our chronic pain GWAS was limited with only one gene - *PIK3C2A –* being identified in both. Identifying an association between both common and rare variants within this gene and pain susceptibility phenotypes suggesting that *PIK3C2A* is likely to be the candidate gene for the GWAS association. *PIK3C2A* encodes an enzyme in the phosphoinositide 3-kinase (PI3-kinase) family which are involved in intracellular protein trafficking, cell survival and the inflammatory response implicating these pathways as potentially important in determining pain susceptibility (54). Several other genes showing the strongest suggestive associations in gene burden-test analysis, despite not overlapping with GWAS loci, are involved in relevant biological mechanisms in pain. For example, *TNFAIP8L1*, otherwise known as TIPE1, is part of a family of immunity regulators, interacting with Rac1 to increase caspase-mediated apoptosis (55). Caspase-mediated apoptosis has previously found to play a critical role in neuropathic and inflammatory pain and could be a potential target for novel pharmacological interventions (56, 57). The burden of rare loss-of-function (LoF) variants within *IGF2BP2* were suggestively associated with chronic pain (*P* = 4.4 × 10^−5^; Supplementary Table 4) and another member of the same gene family, *IGF2BP3*, was prioritised through GWAS (*P* = 1.5 × 10^−8^; Table 3). These insulin-like growth factor 2 mRNA binding proteins may affect the pain pathological process through regulation of RNA metabolism, transcriptional regulation of pain-related genes and release of inflammatory mediators (58–60). The identification of these novel rare variant associations merit further investigation as they may impart larger effect sizes than common variants and highlight important chronic pain pathophysiological mechanisms.

We show a high degree of genetic correlation between our chronic pain phenotypes and specific conventionally defined pain conditions, such as fibromyalgia, migraine, and osteoarthritis, with fibromyalgia demonstrating the strongest significant genetic correlations. Fibromyalgia is a phenotype difficult to define, with a prevalence of up to 2.2% in our study cohort, and has limited well-powered published GWAS studies. This strong genetic correlation with chronic pain phenotypes indicates that the use of our prescription-based phenotyping can potentially identify more fibromyalgia patients than is formally diagnosed and can detect relevant pain related genes that would otherwise be missed using diagnostic criteria. Migraine was found to only have significant genetic correlations with chronic pain duration and susceptibility. This may reflect our definition of pain severity, which is based on opiate use, a class of drugs not used or effective in migraine – hence the pain severity phenotype patients being relatively depleted for migraine sufferers. Osteoarthritis was found to be genetically correlated with chronic pain susceptibility, severity, and duration phenotypes consistent with osteoarthritis being a progressive chronic pain disorder.

Chronic pain is frequently co-morbid with mental health disorders, and we found evidence of shared genetics with psychiatric traits. These findings are consistent to those reported in previous GWAS’ for other pain-related phenotypes (10, 11, 32). In contrast we did not identify genetic correlation of chronic pain with immune disorders, suggesting distinct underlying genetic mechanisms. This does not preclude shared environmental risk factors that may instead explain the frequent comorbidity of pain and immune disorders. Further interrogation of which genes and potential pathways are shared between chronic pain and these conditions will yield better pathophysiological understanding, improved clinical care and could lead to development of therapies to effectively treat both.

The expression and functional enrichment analyses highlight potential underlying mechanisms involved in chronic pain. Using cell-type specific enrichments and mouse phenotype gene-sets, we found novel association of susceptibility to chronic pain requiring opiates with oligodendrocyte progenitor cells (OPCs), gut neurons and peptidergic nociceptors. Chronic pain duration, defined by extended analgesic use, was associated with neuronal and inflammatory cell-types, namely microglia and gut B cells. Interestingly migraine GWAS loci (12) were also enriched in microglia and was correlated with the analgesic use duration phenotype, highlighting modulation of microglia as a therapeutic opportunity in migraine.

Given the lack of other cohorts with sufficient linkage of genetic data with prescription data, additional studies will be required to validate our approach. It is however reassuring that our study re-identified many previously associated chronic pain loci as well as novel loci. Utilisation of a UK Biobank reference panel in LDSC analysis may have introduced biases in the estimation of LD scores given that our study also included Finnish individuals (61, 62). Specifically, genetic variation, allele frequencies, and LD patterns can differ significantly between populations of varying ancestries (63). These differences can lead to inaccuracies in LDSC and impact the reliability and interpretation of heritability estimates and genetic correlations derived from the analysis. Therefore, it’s important to be cautious when generalising findings beyond the populations represented by the European reference panel. Sensitivity analyses using diverse reference panels or population-specific LD scores will help validate and strengthen the results from this study.

Collectively, our advances in pain phenotyping paired with the integration of GWAS, rare-variant gene-based tests, and TWAS highlights novel genes implicated in pain that may have high public significance given current unmet medical need for chronic pain and challenges with existing analgesic therapies. Pathway enrichment highlights neuronal guidance pathways (containing genes including *DCC*, *SEMA3B* and *SEMA3F*), the endolysosomal and post-synaptic recycling endosomal pathways. This should help direct future cellular and *in vivo* studies to elucidate pain mechanisms. A similar prescription-based phenotyping approach could be taken for other heterogeneous traits which are also poorly diagnosed in the clinic, leading to better understanding of the underlying pathophysiology of disease and eventual potential identification of novel therapeutic targets.

## Methods

### Overview of analyses

An overview of the analyses is outlined in Figure 6. We defined 11 novel chronic pain phenotypes (Supplementary Table 1) using prescription data from electronic health records in UK Biobank and FinnGen (Supplementary Table 2). We then performed GWAS followed by meta-analysis including sex-stratified analyses. We also performed gene-based tests using UK Biobank whole genome sequencing (WGS) data. Downstream analysis included prioritisation of candidate genes for the GWAS associations, estimated heritability, genetic correlations, alongside functional enrichment analyses including pathways, mouse phenotypes and cell-types.

**Figure 6:**
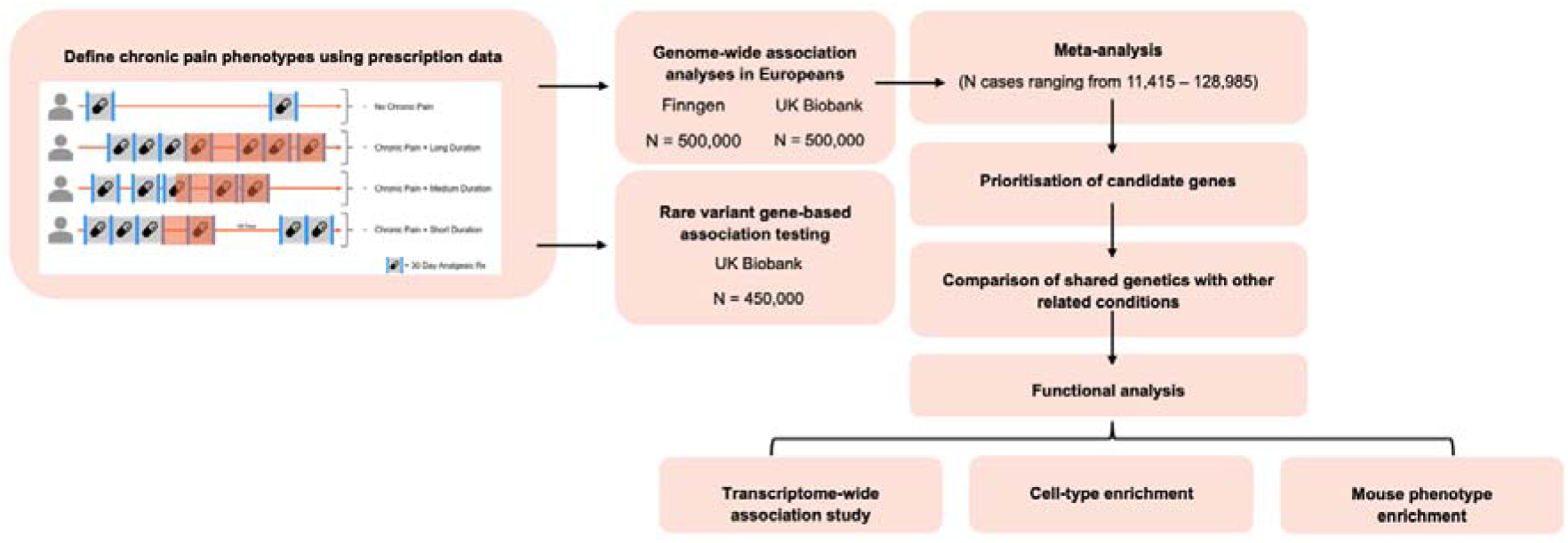
Overview of the analyses performed to identify novel genes and pathways implicated in chronic pain. Briefly, we used analgesic use prescription data in electronic healthy records to define 11 novel chronic pain phenotypes in UK Biobank and FinnGen. Using these 11 newly defined phenotypes, we performed GWAS in both cohorts, followed by meta-analyses to identify common variants associated with chronic pain. The most likely causal genes in each GWAS associated locus were prioritised based on a series of complementary computational approaches, including nearest gene, e/pQTL colocalisations and the presence of high or moderate impact functional variants in the credible set. We investigated the genetic overlap with other pain-related conditions as well as other conditions comorbid with pain, such as psychiatric and immune-related. The genetic associations were then integrated with orthogonal types of data, including cell-type gene expression data, pathway enrichment, and mouse phenotype enrichment, to provide additional insights into underlying biology and pathophysiology of pain.

### UK Biobank

The UK Biobank is large prospective population-based population with over 500,000 participants aged 40-69 recruited across the UK between 2006-2010 (64). UK Biobank contains extensive phenotypic and genotypic details about participants including questionnaire data, physical measures, samples, accelerometery, genome-wide genotyping and longitudinal follow-up for a range of health-related outcomes. Primary care data, providing structured longitudinal and prescription data, for ∼230,000 participants was made available in 2019 and was used for phenotype definition. UK Biobank obtained informed consent from all participants. Given that the incomplete linkage of primary care data in the UK Biobank, we required any control populations to only be selected from subset of ∼230,000 UKB participants with primary care linkage.

### FinnGen

The FinnGen study is a large-scale genomics initiative that has analyzed over 500,000 Finnish biobank samples and correlated genetic variation with health data to understand disease mechanisms and predispositions (65). The project is a collaboration between research organisations and biobanks within Finland and international industry partners. FinnGen provides nationwide longitudinal digital health record data from Finnish health registries collected since 1969. The registries contain coverage on all health-related events, including hospitalizations, prescription drug purchases, medical procedures or deaths.

### Phenotype Definitions

We constructed 11 binary phenotypes describing chronic pain, pain severity, and pain duration (Supplementary Table 1). These phenotypes cover both comparisons to a ‘healthy control’ population (e.g. individuals that did not meet our core definition of chronic pain); and, comparisons within the chronic pain population.

A clinician reviewed a comprehensive list of analgesics prescribed within both UK Biobank and FinnGen. Analgesics were additionally annotated as weak analgesics (e.g. paracetamol, non-steroidal anti-inflammatory drugs (NSAIDs), selective COX inhibitors, triptans and weak opioids) or strong opioids (e.g. buprenorphine, diamorphine, fentanyl, hydromorphone, meptazinol, methadone, morphine, oxycodone, pethidine, tapentadol & tramadol). We assumed that each analgesic prescription is for 30 days duration. While it is possible that actual prescription medication use may not encompass the full 30-day period, our study focused on the intervals between prescriptions to identify both uninterrupted analgesic use and to identify cessation of treatment rather than focusing on the duration of a particular prescription.

The ‘Chronic Pain’ case population comprised of individuals aged ≥18 years with evidence of chronic pain based on at least one period of analgesic prescriptions ≥90 days in duration. A ‘period of analgesic prescriptions’ was defined when: 1) the interval between the date of first analgesic prescription and the date of the last analgesic prescription is ≥90 days duration; and, 2) there is less than a 45 day break between the start of each prescription (assuming a 30-day prescription duration +/- 15 day buffer between prescriptions). The index date for chronic pain was defined as 90 days after the date of first analgesic prescription.

Total analgesic duration (‘Time on Analgesics’) for each patient was quantified by summing the duration of each analgesic prescribing period. Individuals were considered to have ceased analgesic treatment (i.e. entered remission or successfully adopted an analgesic free treatment approach) when a break in analgesic prescription issue dates exceeded 180 days. We defined the time-to-cessation (‘Cessation Time’) for each patient as the number of days from the chronic pain index date to the last analgesic issue date prior to commencement of a period of >180 days without an analgesic prescription. For both ‘Time on Analgesics’ and ‘Cessation Time’, we divided the chronic pain population into quartiles. Our phenotypes then compared: the highest quartile (Q4) to the lowest quartile (Q1) within the chronic pain population; the highest quartile (Q4) to all other quartiles (Q1-Q3) within the chronic pain population; and, the highest quartile (Q4) to healthy controls.

### Genotyping and Imputation

#### UK Biobank

Genotyping, quality control, and imputation was performed centrally by UK Biobank as described by Bycroft *et al.* (66) with a total of 97 million variants (autosomes and X chromosome). We filtered the genotype data to only include variants with minor allele count ≥ 25, imputation quality score (INFO) ≥ 0.3, and Hardy-Weinberg equilibrium exact test *P* value > 1 × 10^−12^. We required a maximum per-variant and per-sample missing call rate <0.01. The final target dataset included 42 million single nucleotide polymorphisms (SNPs) after excluding all duplicate SNPs. Only individuals of European ancestry as defined by the Pan UKBB (https://pan.ukbb.broadinstitute.org/) were included in analyses. The Pan UKBB ancestry assignments are available for download through the UK Biobank portal as Return 2442 (https://biobank.ctsu.ox.ac.uk/showcase/dset.cgi?id=2442). Details about the QC process from the Pan UKBB can be found here including determination of ancestry groups (https://pan.ukbb.broadinstitute.org/docs/qc/index.html)). Genetic sex was defined according to UK Biobank Field 22001 (https://biobank.ndph.ox.ac.uk/showcase/field.cgi?id=22001).

#### FinnGen

We used genotype data from FinnGen Release 11 (R11). FinnGen individuals were genotyped with Illumina and Affymetrix chip arrays (Illumina Inc., San Diego, and Thermo Fisher Scientific, Santa Clara, CA, USA). Chip genotype data were imputed using the population-specific SISu v4.2 imputation reference panel of 8,554 whole genomes. Merged imputed genotype data is composed of 116 data sets that include samples from multiple cohorts. In sample-wise quality control steps, individuals with ambiguous gender, high genotype missingness (>5%), excess heterozygosity (+-4SD) and non-Finnish ancestry were excluded. In variant-wise quality control steps, variants with high missingness (>2%), low HWE *P* value (<1 × 10^−6^) and low minor allele count (MAC < 3) were excluded. The final dataset included 21 million SNP for n=473,681 individuals.

### Association analyses and risk loci definition

GWAS analyses were performed in both UK Biobank and FinnGen adjusting for age at chronic pain (for analyses within the chronic pain population) or age at baseline assessment (for analyses comparing chronic pain to healthy controls), genetic sex, genotype chip, and the first 10 principal components. Logistic regression using an additive genetic model was run in the Regenie software (67). We then performed an inverse-variance weighted fixed effects meta-analyses.

To define independent risk loci, the variant with the lowest *P* value across the genome was defined as an index variant and a 1 mega base (Mb) locus centered at the index variant was defined. This was repeated until no further variants reaching genome-wide significance (*P* values < 5 × 10^−8^) remained.

To assess the overlap of genetic associations across the 11 pain phenotypes, unique loci were identified using an arbitrary 1 Mb distance of one index variant from another index variant. The VennDiagram (68) and the UpSetR (69) packages were used in R (70) to determine the overlap of unique loci across pain phenotypes (chronic pain, pain duration and pain severity) and across sexes.

To determine whether genetic associations identified in our meta-analyses of our 11 chronic pain phenotypes were novel, we compared our genetic findings to those recently reported in the literature for pain or pain-related conditions, including pain susceptibility (10), general pain factor (31), pain intensity (11) and previously reported pain genes reviewed by Li *et al.*(7) (Supplementary Table 16). We also compared to previously reported loci associated with two additional pain-related disorders: osteoarthritis (9) and migraine (12) (Supplementary Table 16). To align with the human assembly used in this study (GrCh38), the chromosome and position of the loci from each study were converted from GrCh37 to GrCh38 using UCSC’s LiftOver tool (71).

### Rare variant burden testing

Rare-variant gene-based burden analyses were performed using WGS data from the UK Biobank. The WGS methods are described in detail in Li *et al.* (72). Briefly, 490,640 UK Biobank participants were sequenced to an average depth of 32.5X using the Illumina NovaSeq 6000 platform. Variants were jointly called using Graphtyper, which resulted in 1,037,556,156 and 101,188,713 high quality (AAscore < 0.5 and < 5 duplicate inconsistencies) SNPs and indels respectively. We further processed the jointly called genotype data in Hail v0.2, where multi-allelic sites were first split and normalized. Variants were then filtered based on low allelic balance (ABHet < 0.175, ABHom < 0.9), low quality-by-depth normalized score (QD < 6), low phred-scaled quality score (QUAL < 10) and high missingness (call rate < 90%).

We defined a cohort of European ancestry individuals to be that most resembled the NFE (non-Finnish European) population as labelled in the gnomAD v3.1 dataset (73). Variant loadings for 76,399 high-quality ancestry-informative variants from gnomAD were used to project the first 16 principal components onto all UK Biobank WGS samples. A random forest classifier trained on the nine gnomAD ancestry labels was then used to assign individuals to ancestry groups (probability > 0.9). In total, 458,855 individuals of European ancestry were taken forward for analysis.

Variants were annotated using Ensembl’s VEP tool (74) and putative loss of function (LoF) variants were annotated using the LoFTEE plugin.

For burden testing, we defined five masks using variants with minor allele frequency (MAF) < 0.001: 1) LoFHC (LoFTEE high confidence), 2) All LoF (LoFTEE high and low confidence), 3) All LoF and DeleteriousMissense (combination of mask 2 and SIFT or Polyphen prediction being deleterious or probably damaging respectively), and 4) All LoF & Missense (combination of mask 2 and any variant annotated as a missense by VEP), and 5) All Missense (any VEP annotated missense variant). For each phenotype, gene-based burden testing was performed using the SKATO method as implemented in Regenie v3.2.5 (67) on the same phenotypes as the UK Biobank GWAS.

### SNP-based heritability and Genetic correlations

LD score regression (LDSC) (75) was used to estimate per-trait SNP-based heritability (h^2^) and pairwise genetic correlation (r_g_) between the novel chronic pain phenotypes and GWAS meta-analyses that focused on pain traits (migraine and osteoarthritis), psychiatric conditions (bipolar disorder, depression, anxiety, post-traumatic stress disorder (PTSD)), and immune-related conditions (rheumatoid arthritis, Crohn’s disease, inflammatory bowel disease (IBD)) (Supplementary Table 17).

LD scores used in the analyses were derived using LDSC from 10000 unrelated non-Finnish European individuals from UK Biobank with available whole genome sequencing (WGS) data. LD scores were estimated by collapsing common (MAF > 0.01) HapMap3 variants within 1Mb windows to comprehensively capture common variant tagging. Variants included in the analyses were restricted to well-imputed HapMap3 variants, per method recommendations.

For some traits, particularly those which were sex-stratified, low statistical power led to inaccurate estimations of SNP-based heritability (h² ∼ 0.01 or h^2^ < 0). This meant that pairwise genetic correlation could not be estimated for a subset of trait combinations.

### Prioritisation of most likely causal genes

To determine the most likely causal gene at each loci, we performed three complementary approaches: 1. ‘Nearest Gene’: we identified the nearest protein-coding gene to the lead variant based on distance to the transcription start site or the gene body; whichever was closer. 2. ‘Coding variant’: we used VEP (74) to annotate variants with their predicted effect and considered any coding variant with high or moderate impact (missense, frameshift, stop gained, start/stop lost, inframe insertion/deletion) within the credible set of the signal with a posterior probability > 0.01 (76) as evidence for prioritisation. 3. ‘Quantitative trait locus (QTL) colocalization’: We performed colocalization of the pain GWAS signals with pQTLs and eQTLs. For eQTLs, we used data from the Genotype-Tissue Expression Consortium (https://gtexportal.org, (22)). For pQTLs, we used data from UK Biobank (77), DeCODE (78) and Fenland (79). For both eQTLs and pQTLs, we considered only those acting in *cis,* defined as genes located < 1 MB from the lead variant. We performed multicolocalization analysis using the *coloc* package (80) in R (70). We determined a significant colocalization if the GWAS signal had a *P* < 5 × 10^−8^, the QTL signal had a *P* < 1 × 10^−5^ and the posterior probability for the signals being shared was greater than 0.8 (H12 > 0.8).

### Transcriptomic Wide Association Analysis

Using the S-PrediXcan software (81), we analysed cis-eQTLs from Bryois *et al.* (21) for eight brain cell-types derived from 373 human brain samples from 215 individuals, as well as tibial nerve eQTLs from the GTEx project (22). We tested the genetic association between chronic pain phenotypes and both cell-type-specific and tibial nerve eQTLs. We identified TWAS significant genes as those that passed multiple testing corrections for the number of genes analysed in the TWAS for each tissue and cell-type. The TWAS process involves two stages. First, gene expression and genotype datasets are integrated using a regression model, assuming linear additive genetic effects. In the second step, the eQTL effect sizes are used to impute gene expression in an independent GWAS dataset, and the association between the trait and imputed gene expression is tested.

### Gene-set cell-type enrichment

We used publicly available datasets from published studies, including the cortex, substantia nigra (16), trigeminal ganglia (17), dorsal root ganglia (18), and gut (18, 19). The cell-types in each single-cell dataset are listed in Supplementary Table 18. The available information for transcriptomic data sets were in two formats: (1) the mean expression for each gene in each cell type and (2) the gene expressions count matrix. If the mean expression for each gene is available, we used to identify specifically expressed genes and the 10% most expressed genes for each cell type are defined as those with the large mean expression. However, if the gene expression matrix is available, we computed t-statistics for each gene to identify the most specifically expressed genes. To compute a t-statistic, each gene (g) was assigned a value of “1” if the sample is expressed in a cell-type and “-1” if otherwise. We define outcome (y) as scaled gene expression and then fit a simple linear regression model of “y” on “g” and compute t-statistics for each gene. This will produce t-statistics for each gene in each cell-type, and the 10% most expressed genes for a given cell-type are defined as those with the largest positive t-statistics.

We used MAGMA (82) (v1.10) software to perform gene-set enrichment analysis using GWAS summary statistics datasets. Using the GWAS summary statistics dataset, MAGMA performs gene-set enrichment in two steps, (1) gene-level association and (2) gene-set enrichment. In the gene-level association, MAGMA averages SNPs *P* values located around the gene while accounting for LD. In the gene-set analysis, the averaged gene-level *P* values are converted to z-scores and used to test whether a gene-set is more enriched in a phenotype compared to genes not in the gene-set. A 25 kilobases (kb) window upstream and downstream on a gene location was used for gene-level analysis to define a gene window. MAGMA was then used to investigate whether the 10% most specifically expressed gene in each cell-type was associated with the phenotypes. A 5% significant threshold was set to determine significant association after Bonferroni correction.

Partitioned LD score (83) regression was used to investigate whether the 10% most specifically expressed genes in a cell-type account for the heritability of the phenotypes. We used a 25 kb window on both sides of a gene coordinate to define a gene region. The Bonferroni corrected *P* values were used to determine the association of a cell-type to a phenotype and a 5% multiple testing cut-off was used as a threshold. The abstract mediation model (AMM) (84) was also used to test whether the 10% most expressed genes of each cell-type mediated the heritability in each trait. AMM estimates the fraction of heritability mediated by the kth-closest gene to each SNP because in the genome some fraction of heritability is mediated by genes that are not the closest. To account for gene contribution, the 50^th^-closest gene was used as the farthest gene from a SNP. Bonferroni corrected *P* values were used to determine the cell-types enriched for the heritability of the phenotypes.

### Gene-set functional characterisation

To characterise mouse phenotypes associated with pain-related cell-types, the top 10% most specifically expressed genes in each cell-type were tested for enrichment in mouse phenotype gene-sets. Mouse genotype-phenotype associations were obtained from the Monarch Initiative (20), filtered for genotypes affecting a single mouse gene with a high-confidence human ortholog as defined by Ensembl Compara (85), and then aggregated to the gene level. Enrichment was tested using the hypergeometric test implemented with the clusterProfiler R package (86) and was relative to the background set of genes detected in each scRNA-seq dataset. Significantly enriched gene-sets were defined as those with >= 2 overlapping genes, gene ratio >= 0.01, and FDR-adjusted *P* value <= 0.05. Cell-types shown to have cell-type-specific genes significantly enriched in pain-related GWAS signals were selected for further study. This included extracting cell-type-specific genes in enriched mouse phenotype gene-sets to test for pain GWAS enrichment.

### Pathway enrichment analysis and protein-protein interactions

We constructed a protein-protein interaction (PPI) network to represent the structure of interactions in cell-types associated with specific phenotypes. To do this, the PPI for the list of genes in each cell-type were selected and prepared for network analysis using the *igraph* (87) package in R (70). The *cluster_louvain* function was used for building the interactions. This clustering generated modules for each cell-type and modules with at least 30 genes were considered as more informative. The structure of the networks was further simplified to get useful information from them by using only genes with at least 10 interactions. Using *topGO* (88), and *rrvgo* (89) packages in R (70) we perform gene ontology enrichment analysis.

## Data Availability

All data produced in the present study are available upon reasonable request to the authors.

## Acknowledgements

We want to acknowledge the participants and investigators of the FinnGen and UK Biobank studies. This research has been conducted using the UK Biobank Resource under Application Numbers 20361, 68574, & 65851. The authors would like to thank Robert Scott, Sanjay Kumar, Carolyn Buser-Doepner, Emma Laing, Lisa Mohamet, and Niall Moore for their support during this project.

