## Supplementary material for "GWAS of Extended Prescription Analgesic Use Identifies Novel Genetic Loci in Chronic Pain": Supplenetary Figures 1-7

Supplementary Figures

**Supplementary Figure 1: Quantile-Quantile (QQ) plot for the novel chronic pain phenotype defined using analgesic use-prescription data.** The observed negative log_10_ *P* values (y-axis) for each SNP are sorted from largest to smallest and plotted against the expected negative log_10_ *P* values (x-axis) from the χ^2^ distribution.

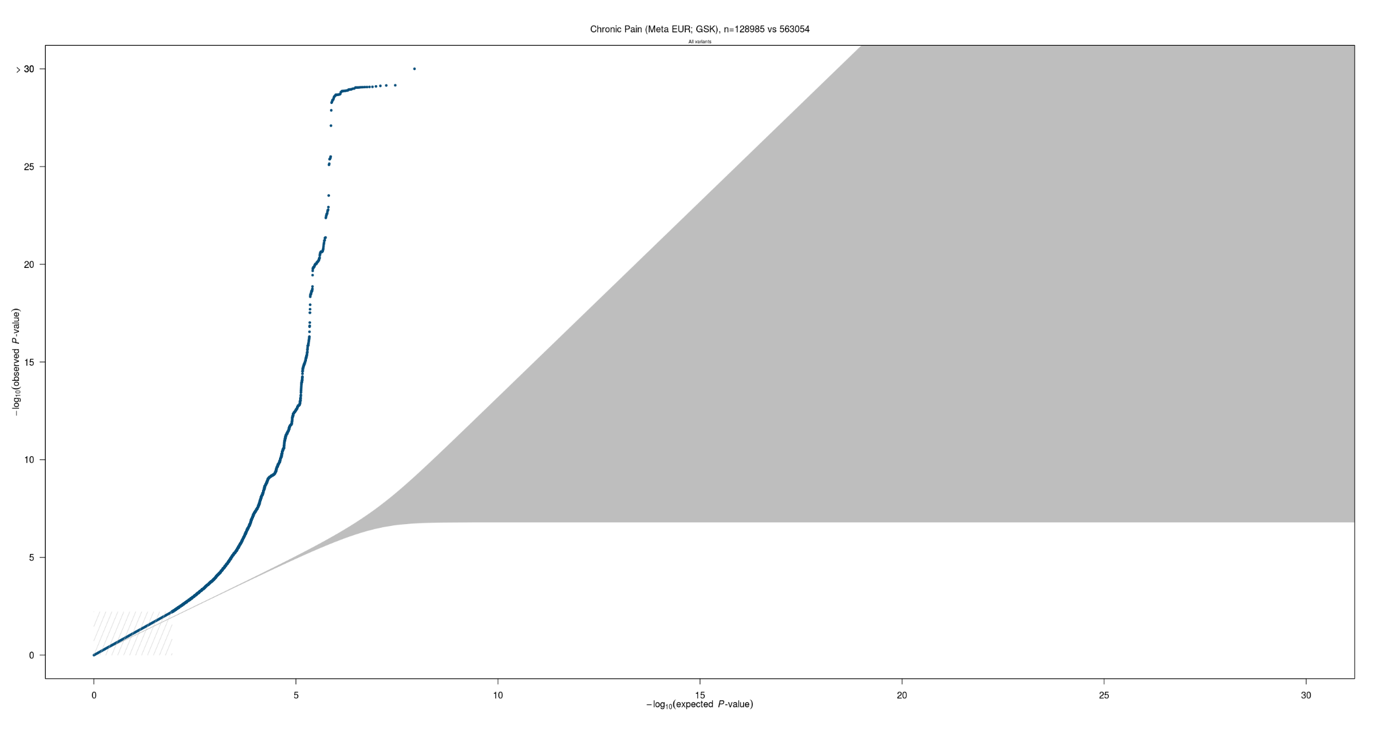

**Supplementary Figure 2: Manhattan and Quantile-Quantile (QQ) plots for the 10 other novel chronic pain phenotypes defined using analgesic use-prescription data.** Each dot on the Manhattan plot (bottom left) represents a single nucleotide polymorphism (SNP). On the x-axis, the SNPs are plotted along the 22 autosomes. Adjacent chromosomes are colored in different shades of blue. The y-axis shows the statistical strength of association from the inverse-variance weighted fixed-effect meta-analysis as the negative log_10_ of the uncorrected *P* value (*P*). The horizontal line is the genome-wide significance threshold after correction for multiple testing (*P* = 5 x 10^-8^). The QQ-plot (top right) of the data shown in the Manhattan plot representing the deviation of observed *P* values (y-axis) from the null hypothesis. The observed negative log_10_ *P* values for each SNP are sorted from largest to smallest and plotted against the expected negative log_10_ *P* values (x-axis) from the χ^2^ distribution. A: Strong Opioid use vs healthy. B: Strong Opioid use for more than 90 days vs healthy. C: Strong Opioid use for > 90 days vs < 90 days. D: Strong Opioid use vs chronic pain patients who have never received strong opioids. E: Time on analgesics Q4 vs Q1-Q3. F: Time on analgesics Q4 vs healthy. G: Time on analgesics Q4 vs Q1. H: Cessation Time Q4 vs Q1-Q3. I: Cessation Time Q4 vs healthy. J: Cessation Time Q4 vs Q1.

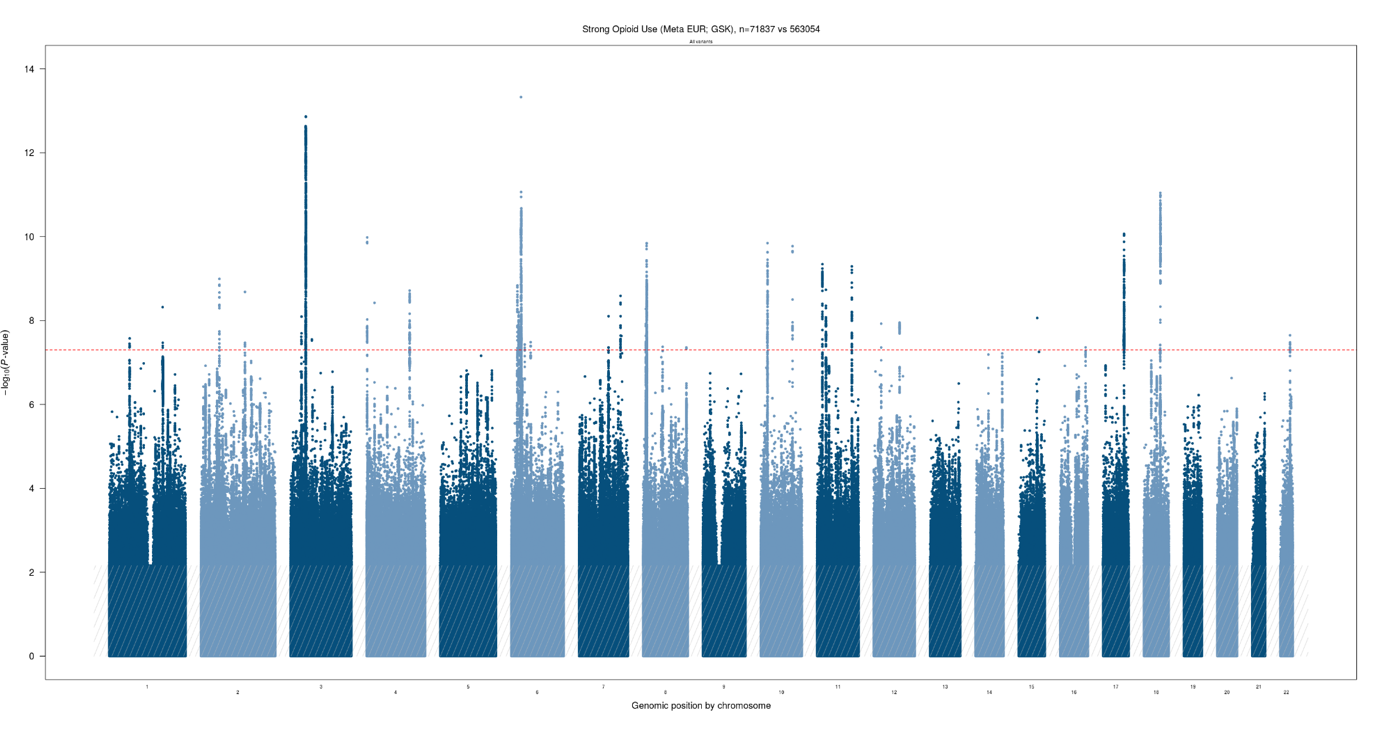

**A**

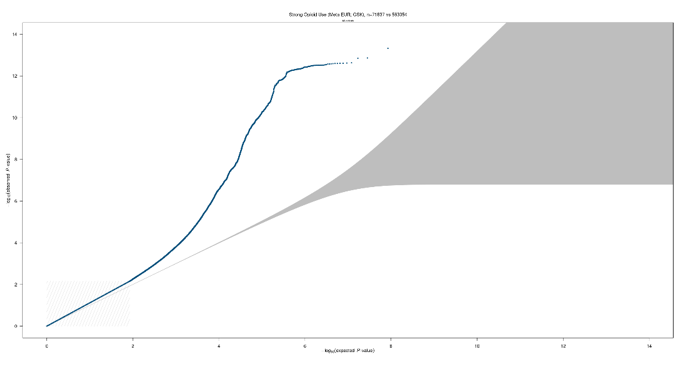

**B**

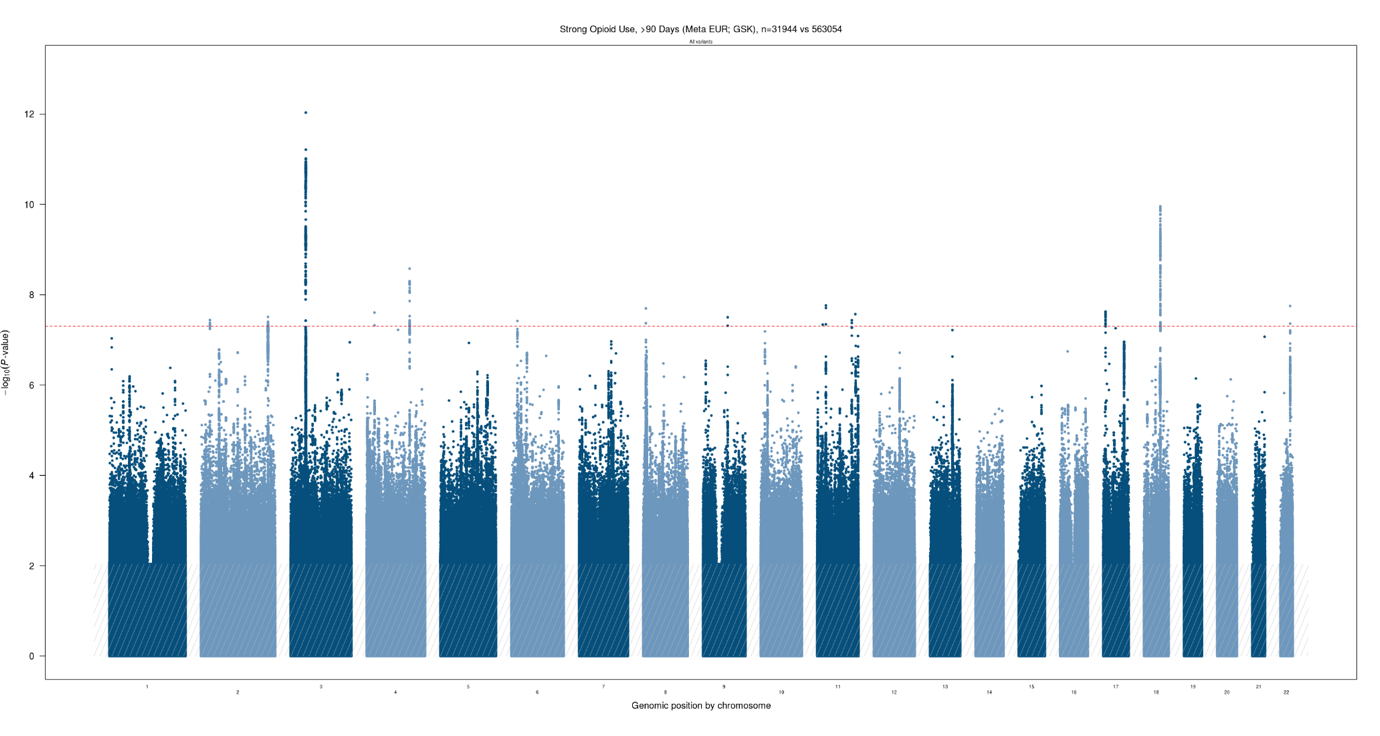

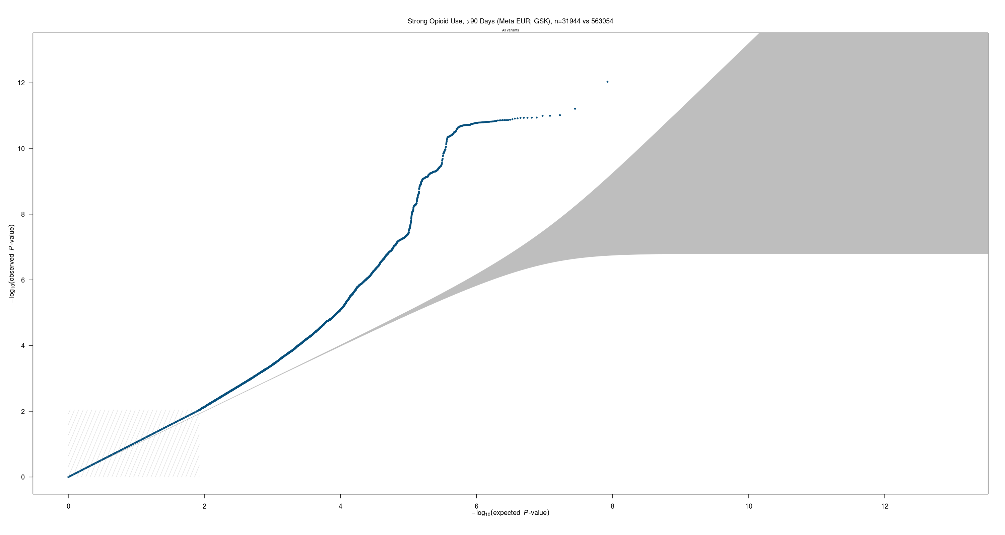

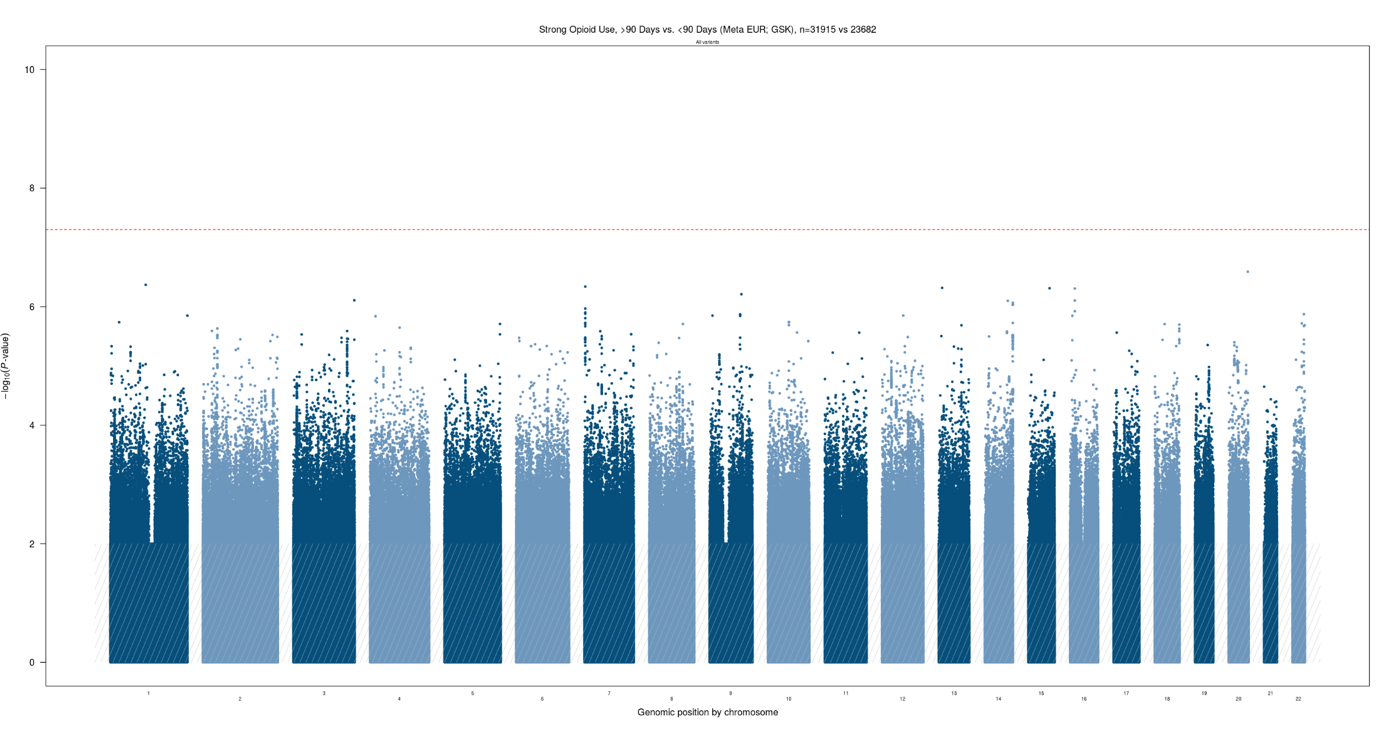

**C**

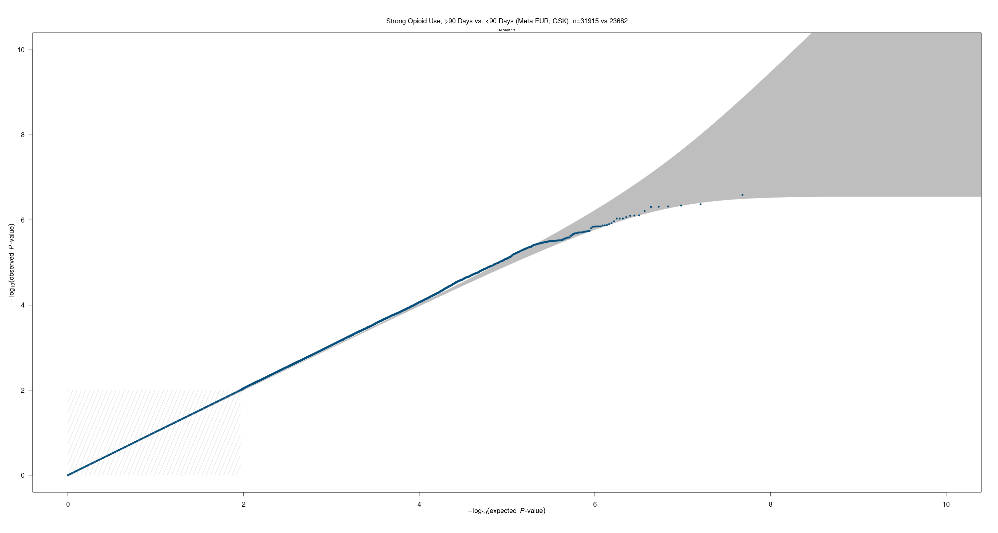

**D**

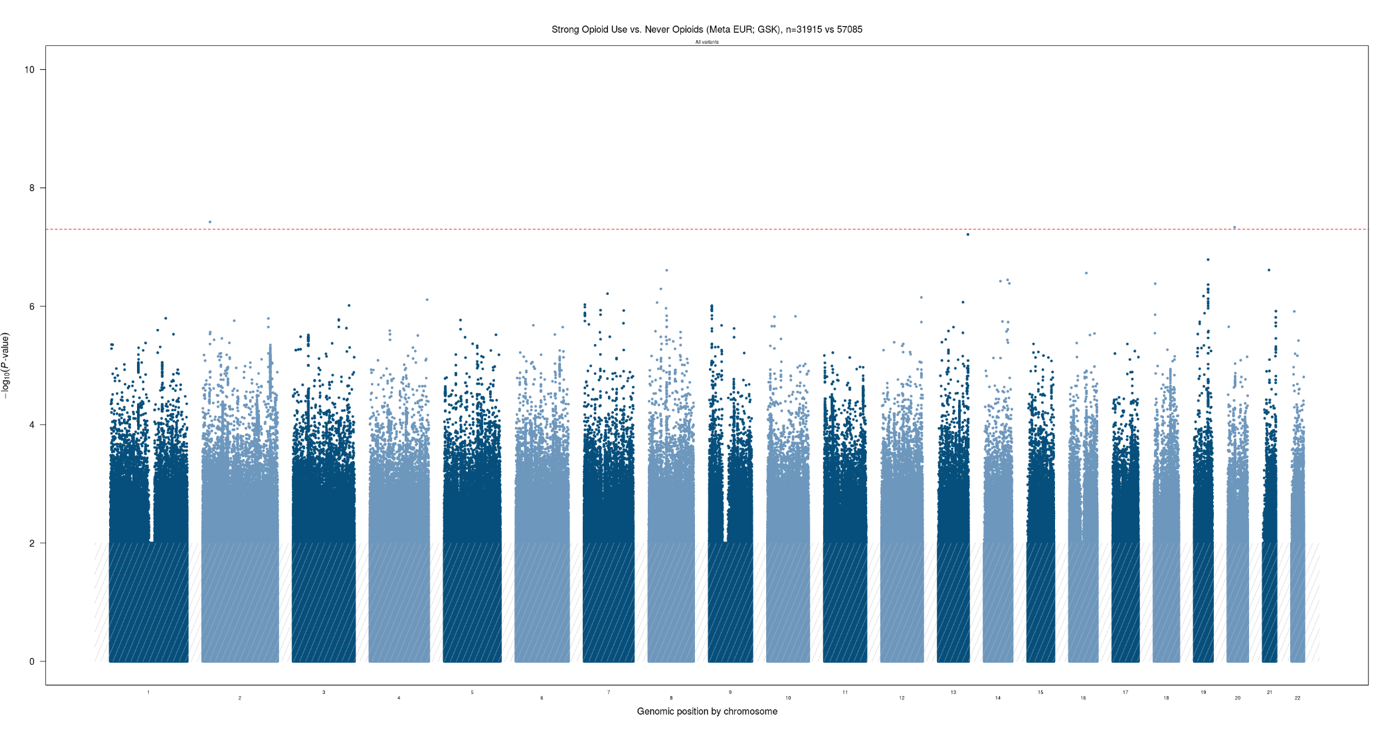

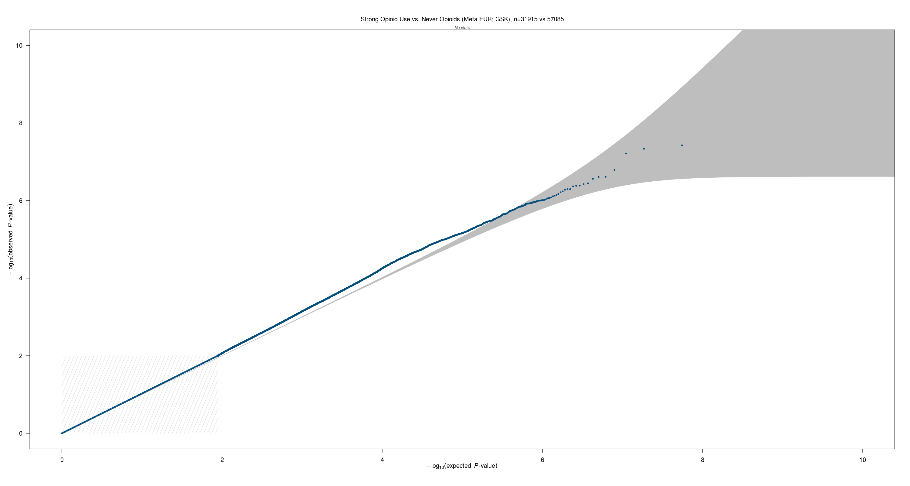

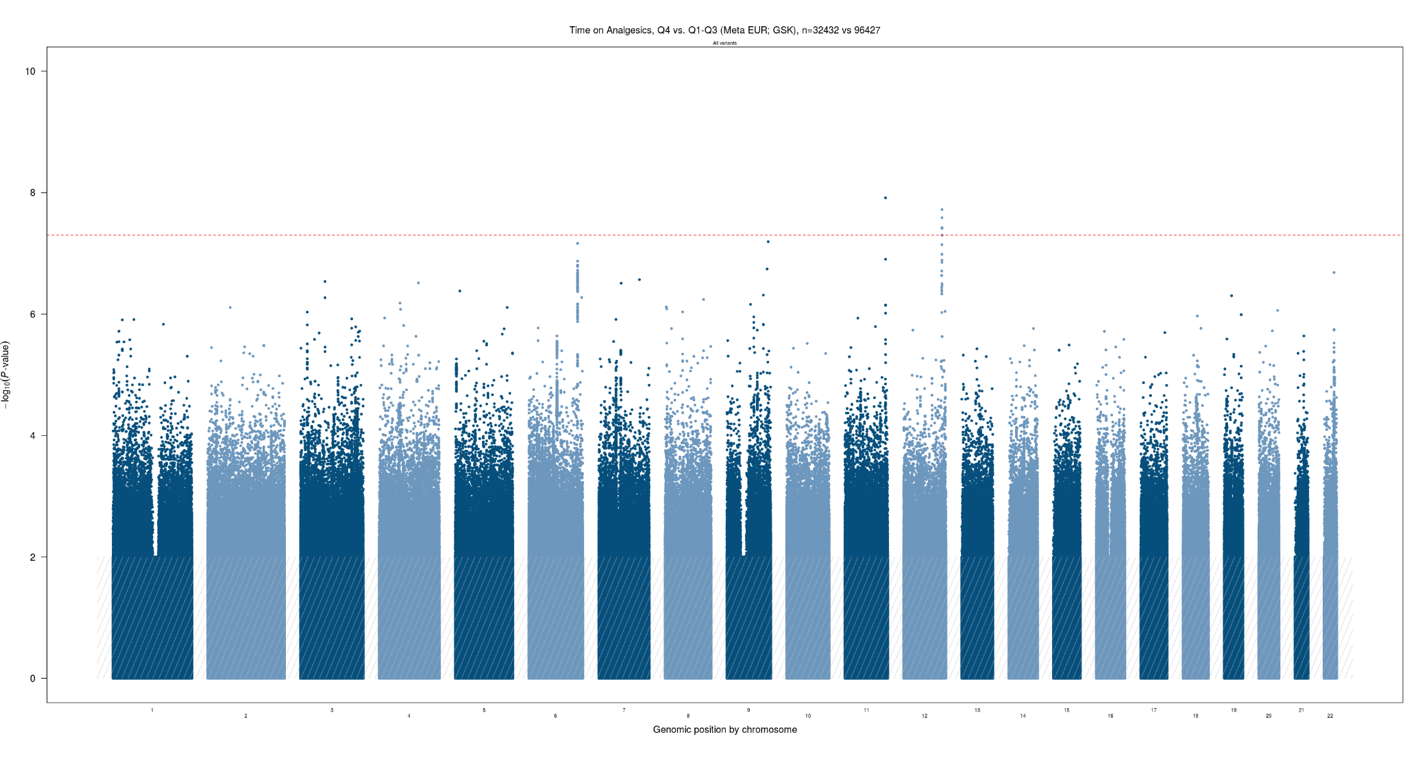

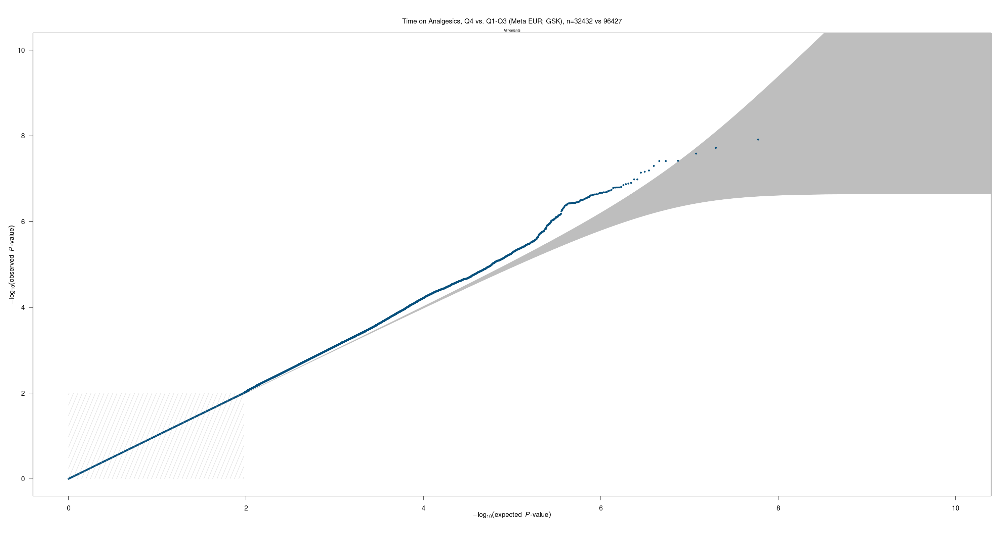

**E**

**F**

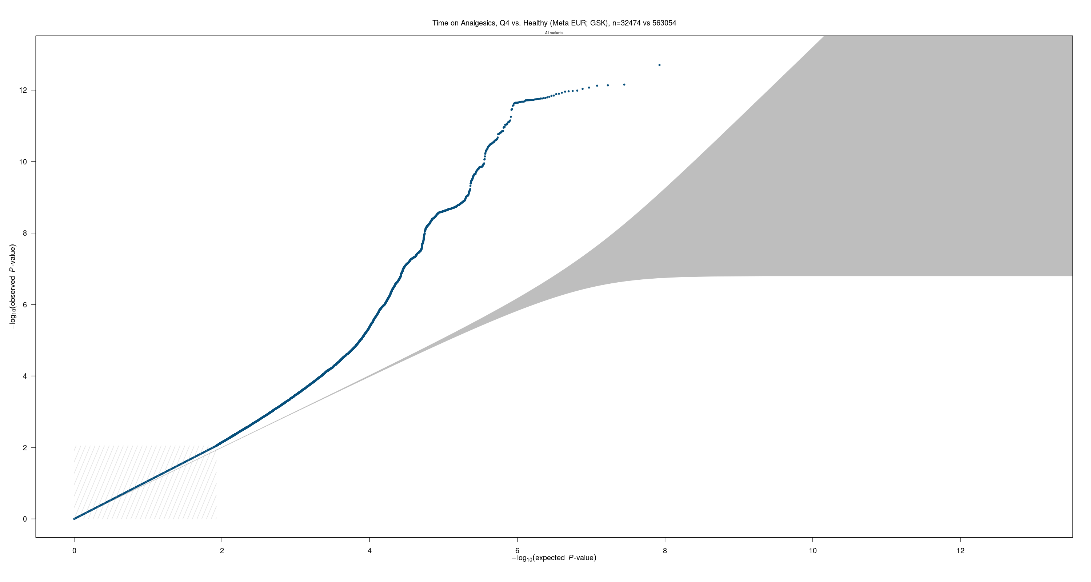

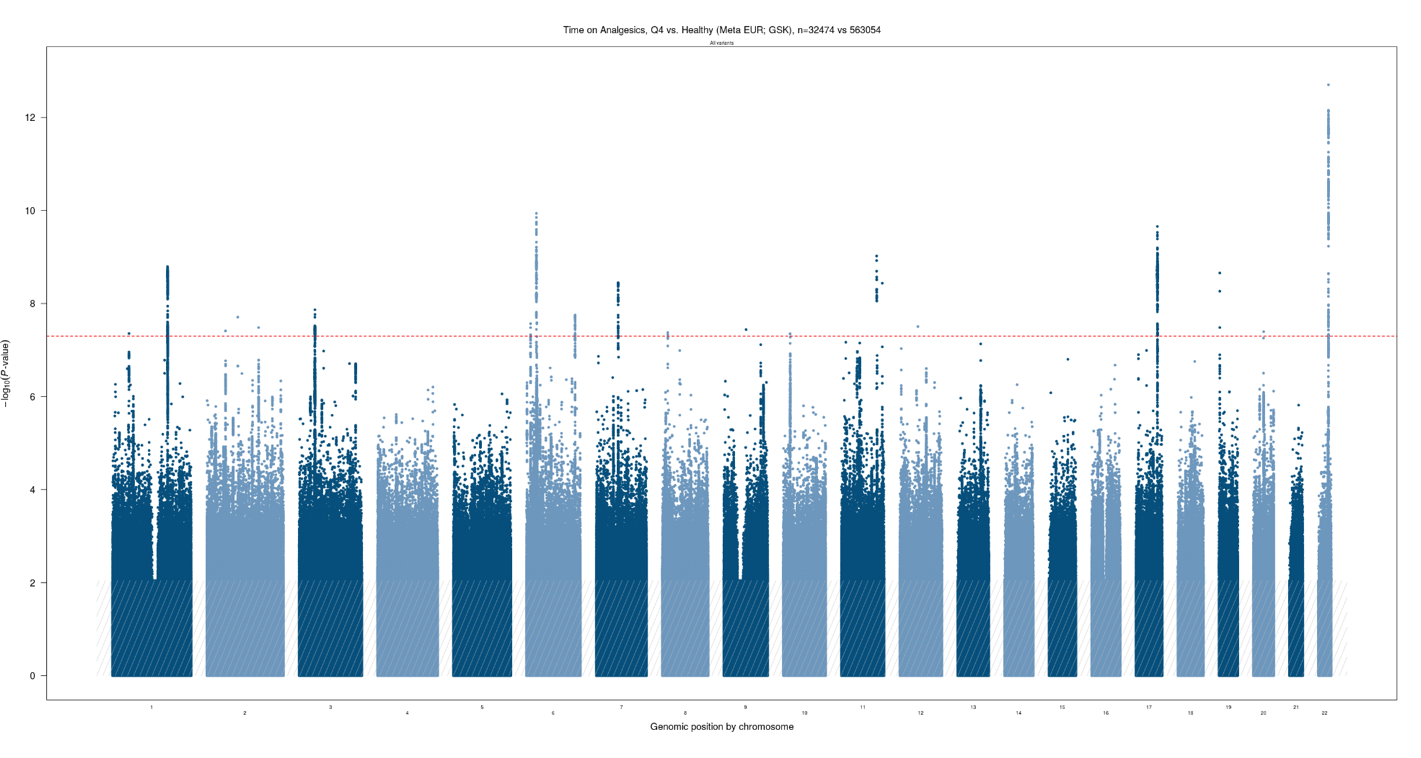

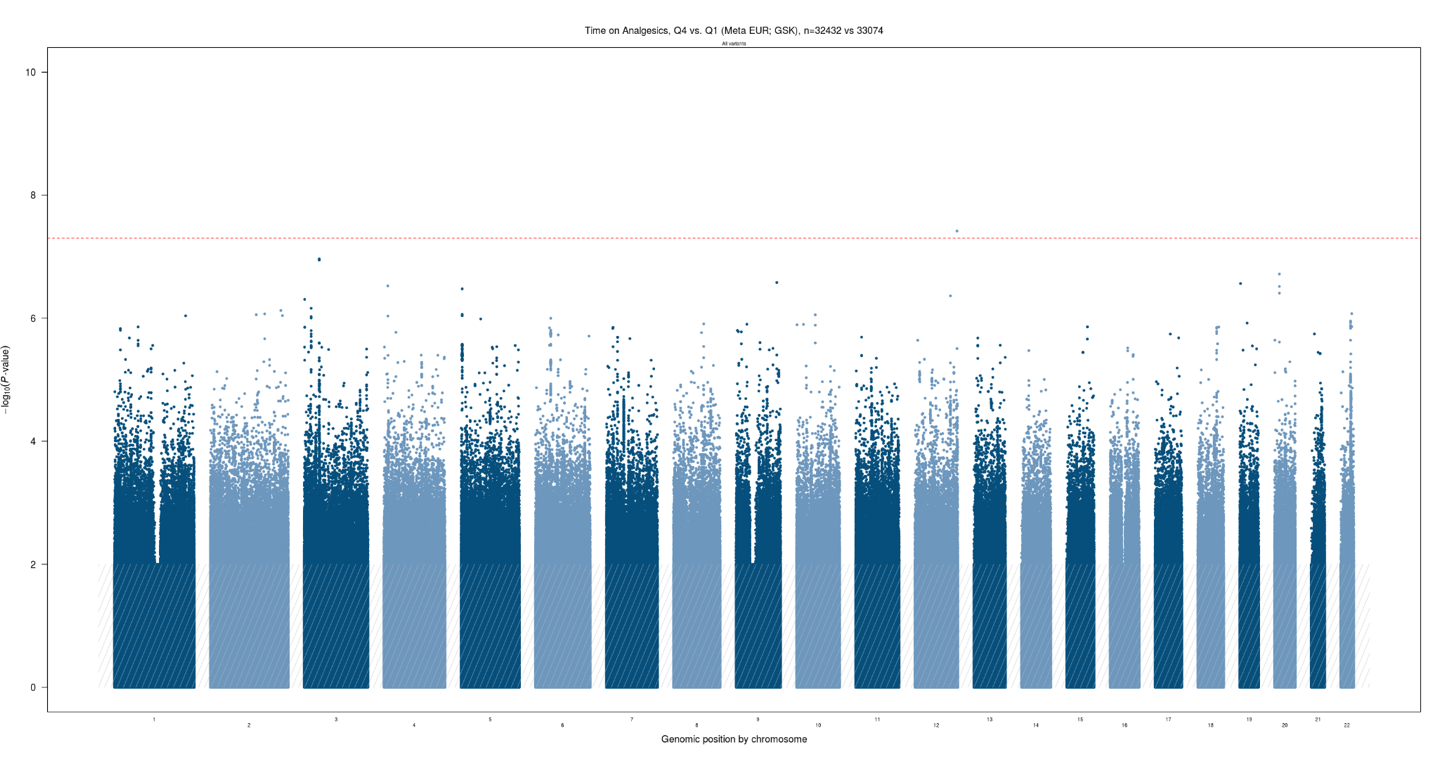

**G**

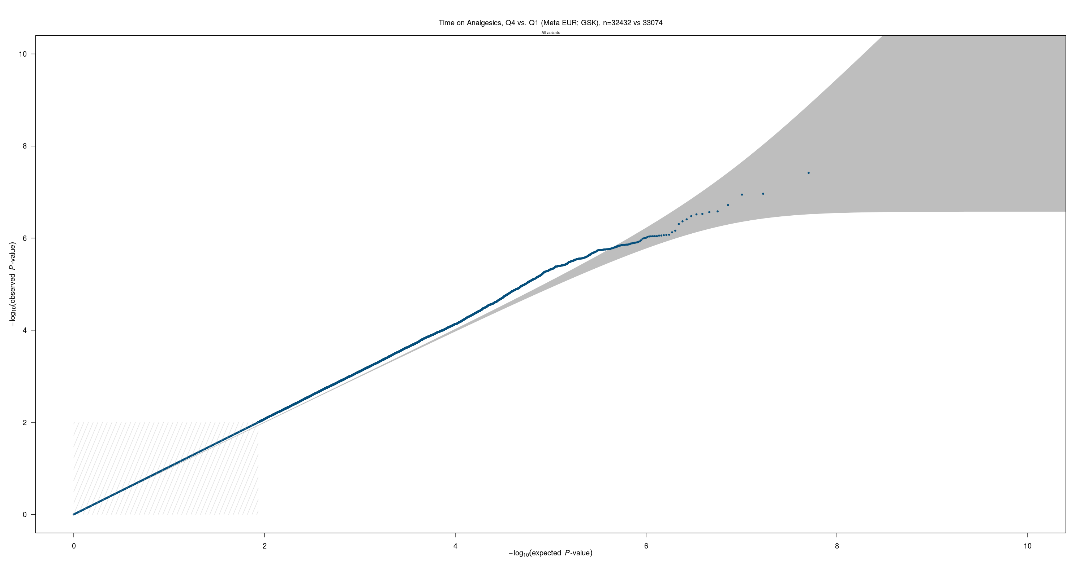

**H**

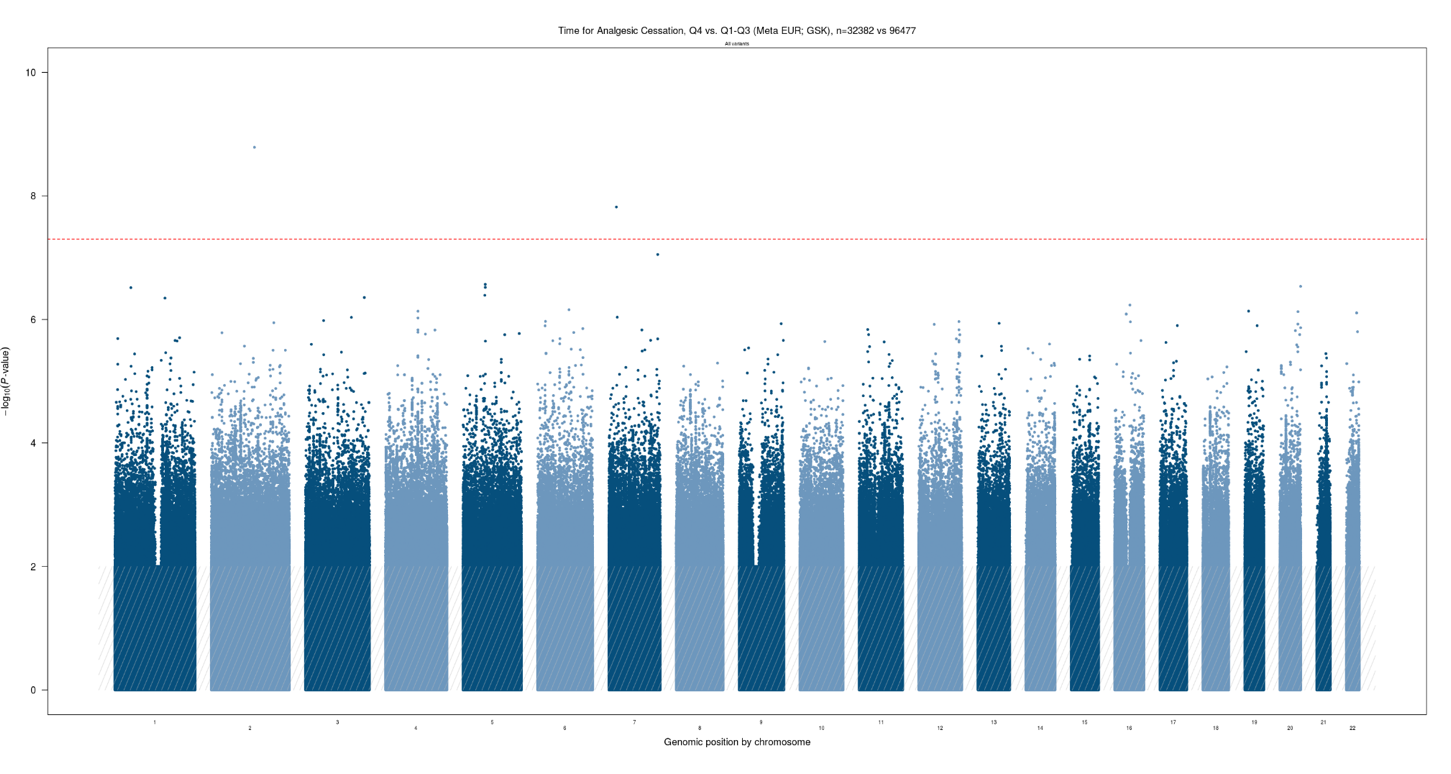

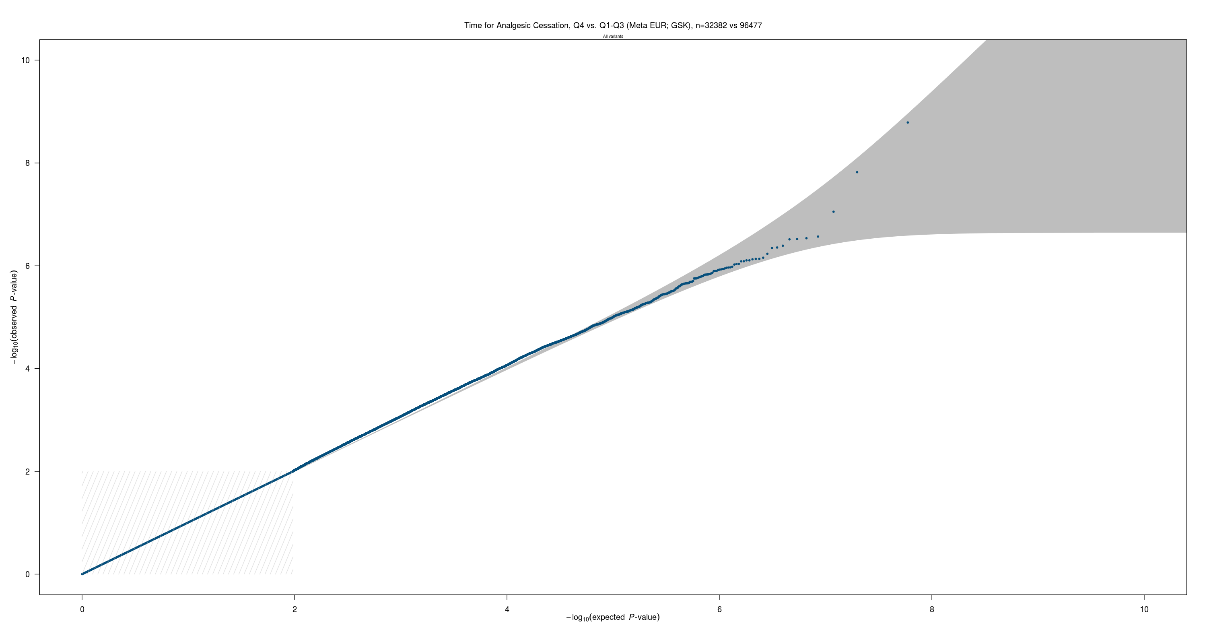

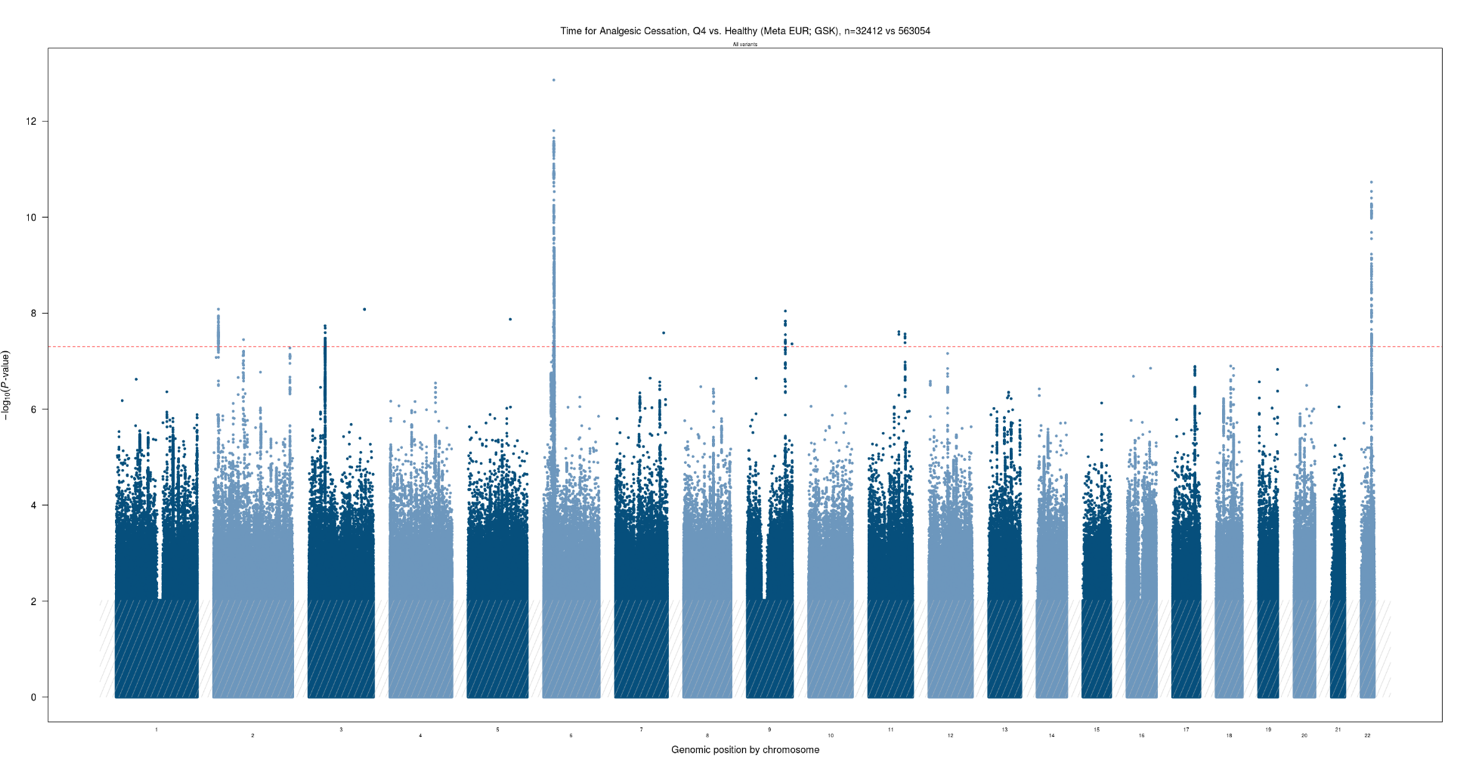

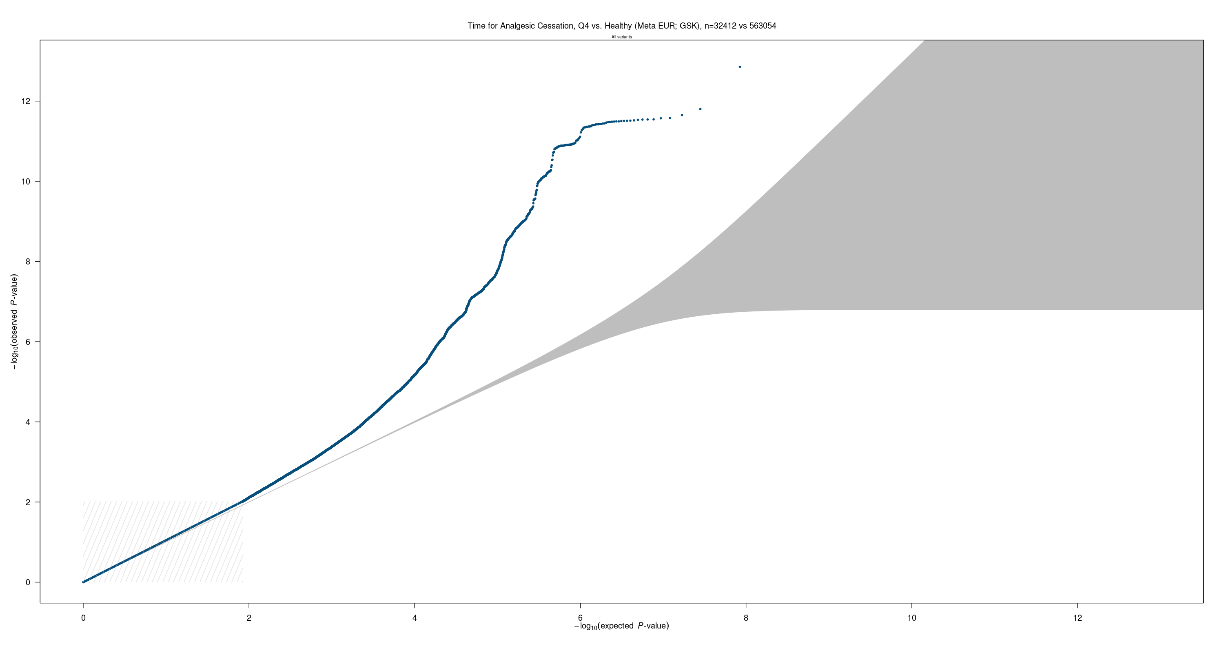

**I**

**J**

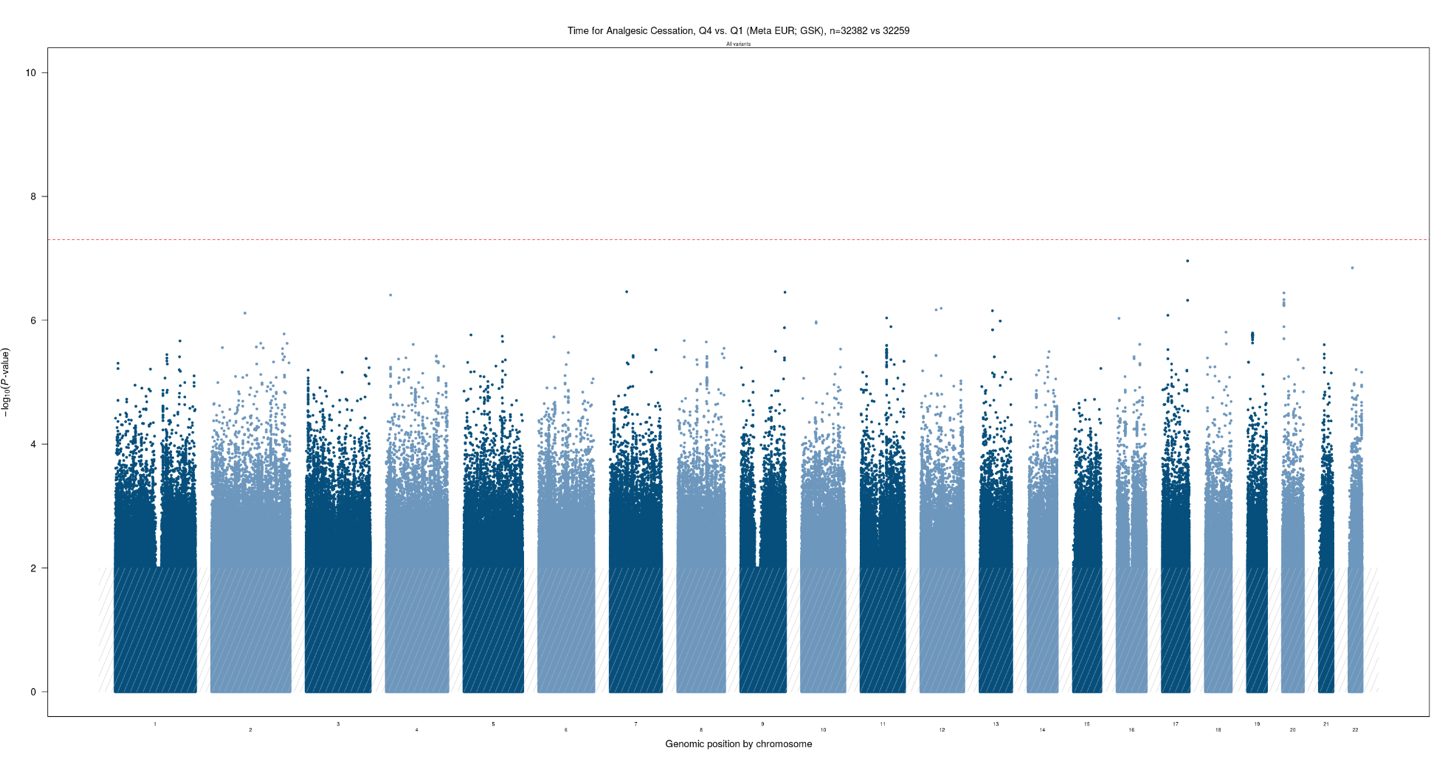

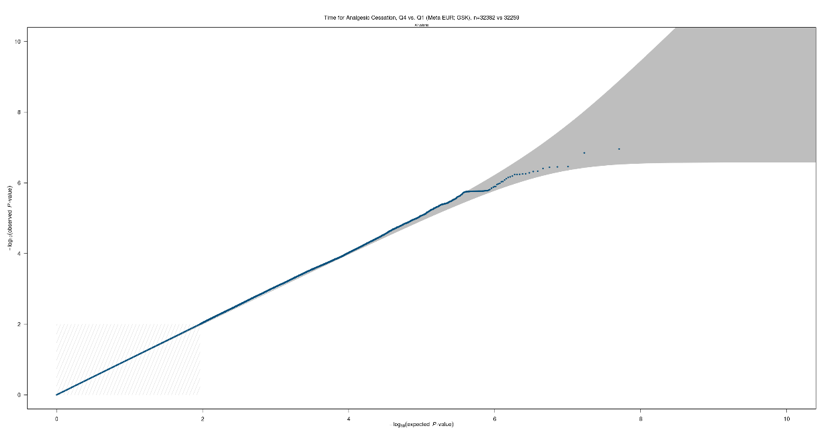

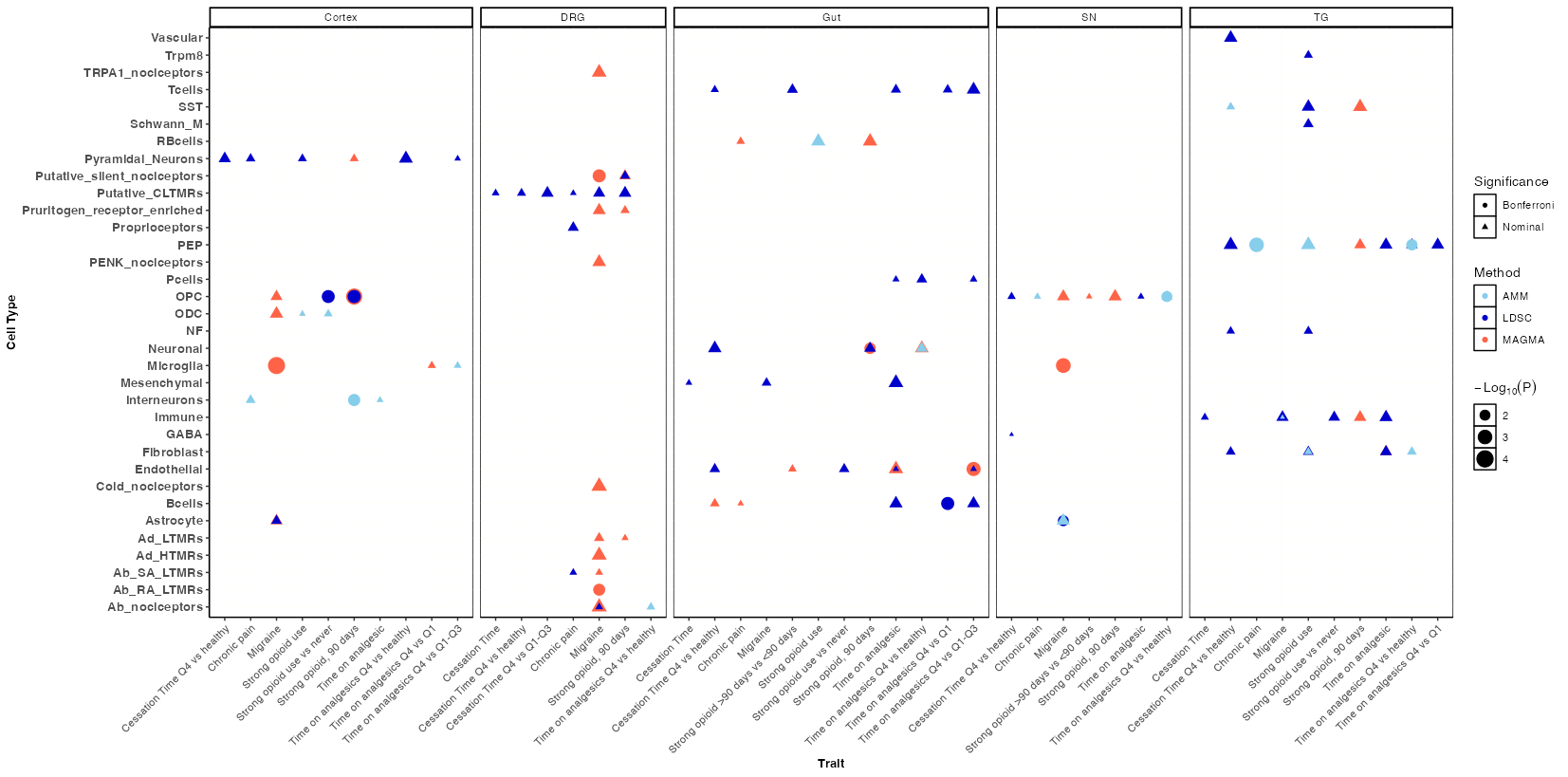
**Supplementary Figure 3: Tissue enrichment analysis**

Cell-types associations with chronic pain phenotypes grouped by tissues. Associations are represented as negative log_10_ (*P* value). The stronger the association the larger the shape. Circles represent associations reaching a Bonferroni adjusted *P* value threshold, whilst triangles represent associations passing nominal significance (*P* < 0.05). The color of the shading represents the statistical method used to identify the association (light blue – AMM, dark blue – LDSC, red – MAGMA). Abbreviations: SST: somatosastin, CLTMRs: , GABA: Gabergic neurons, PEP: Peptidergic nociceptors, RBcells: Red blood cells, OPC: Oligodendrocyte Progenitor cells, ODC: Oligodendrocytes, NF: Neurofilaments, Ad_LTMRs: Ad low-threshold mechanoreceptors Ad_HTMRs: Ad high-threshold mechanoreceptors, Ab_SA_LTMRs: Ab slowly adapting low-threshold mechanoreceptors, Ab_RA_LTMRs: Ab rapidaly adapting low-threshold mechanoreceptors

**Supplementary Figure 4: Heat map of the shared genetic effects between sex-stratified chronic pain phenotypes and other related traits.**

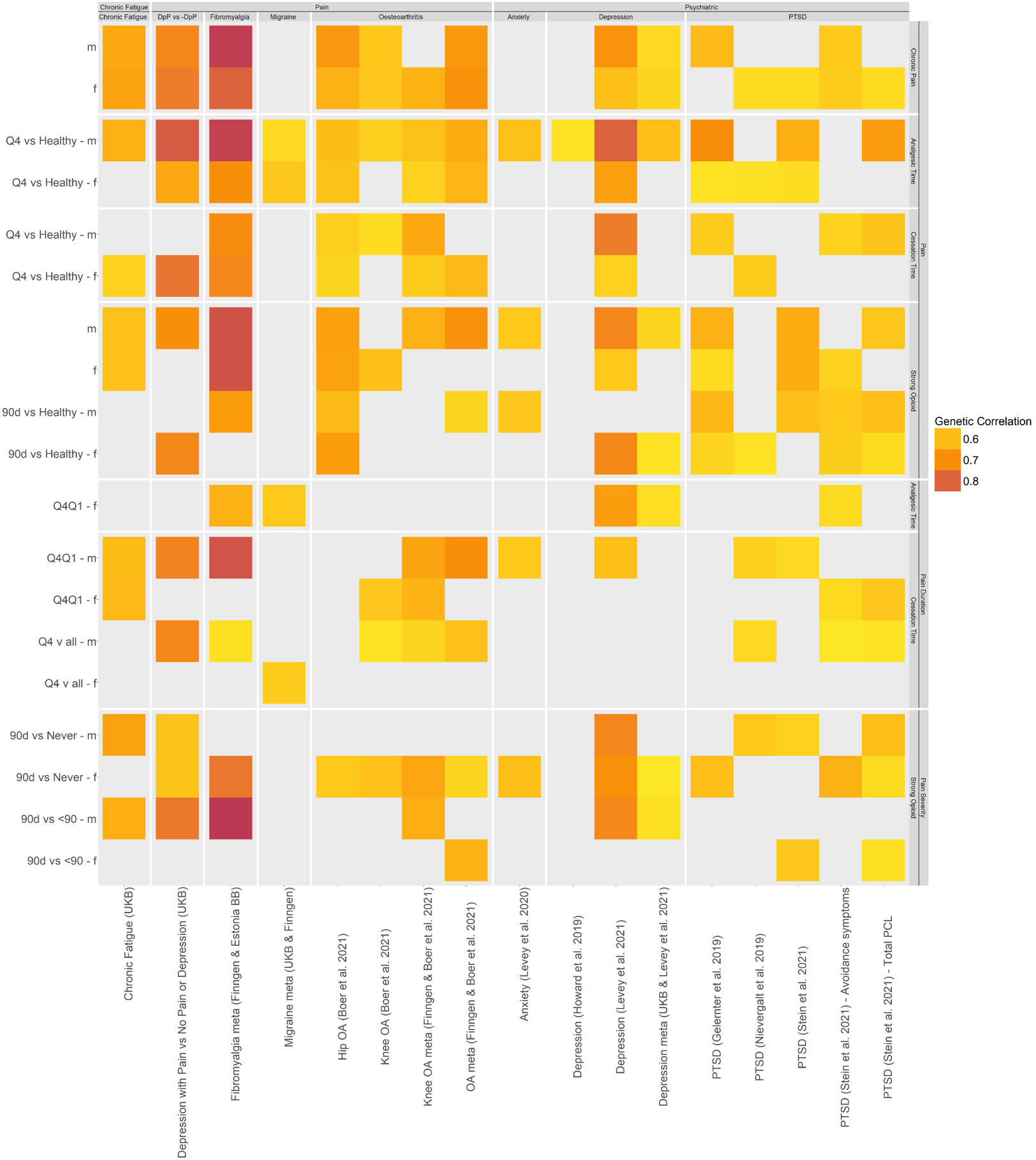

*Heat map shows only those results passing a P<1E-05 and with a genetic correlation > abs(0.5). The y-axis shows the chronic pain phenotypes defined using prescription data clustered by chronic pain, analgesic time, cessation time and strong opioid use. The x-axis shows the related phenotypes clustered by pain, and psychiatric. There were no significant genetic correlations between any chronic pain phenotype defined using prescription data and immune conditions. The darker the red shading the stronger the genetic correlation. Abbreviations: DpP: depression with chronic pain, DpP: No depression or chronic pain.*

**Supplementary Figure 5: Regional association plot of the SEMA3F locus with pain susceptibility (chronic pain vs healthy) and corresponding colocalization plot for the locus.**

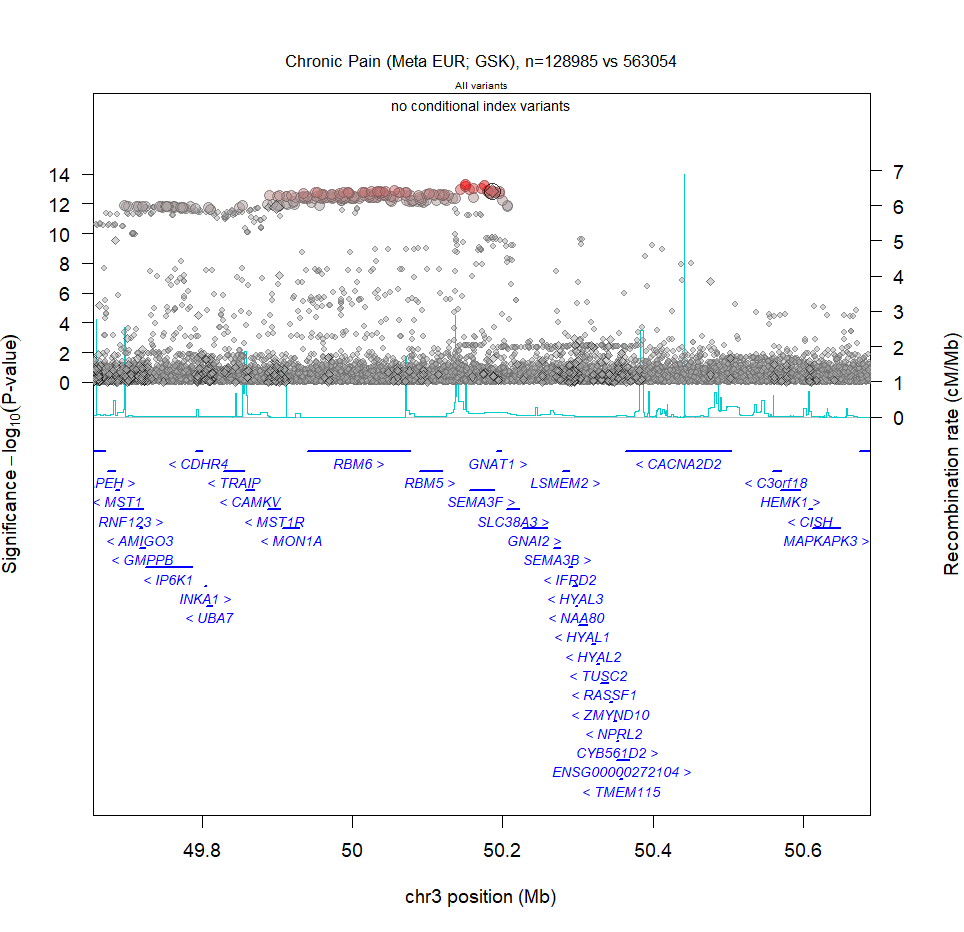

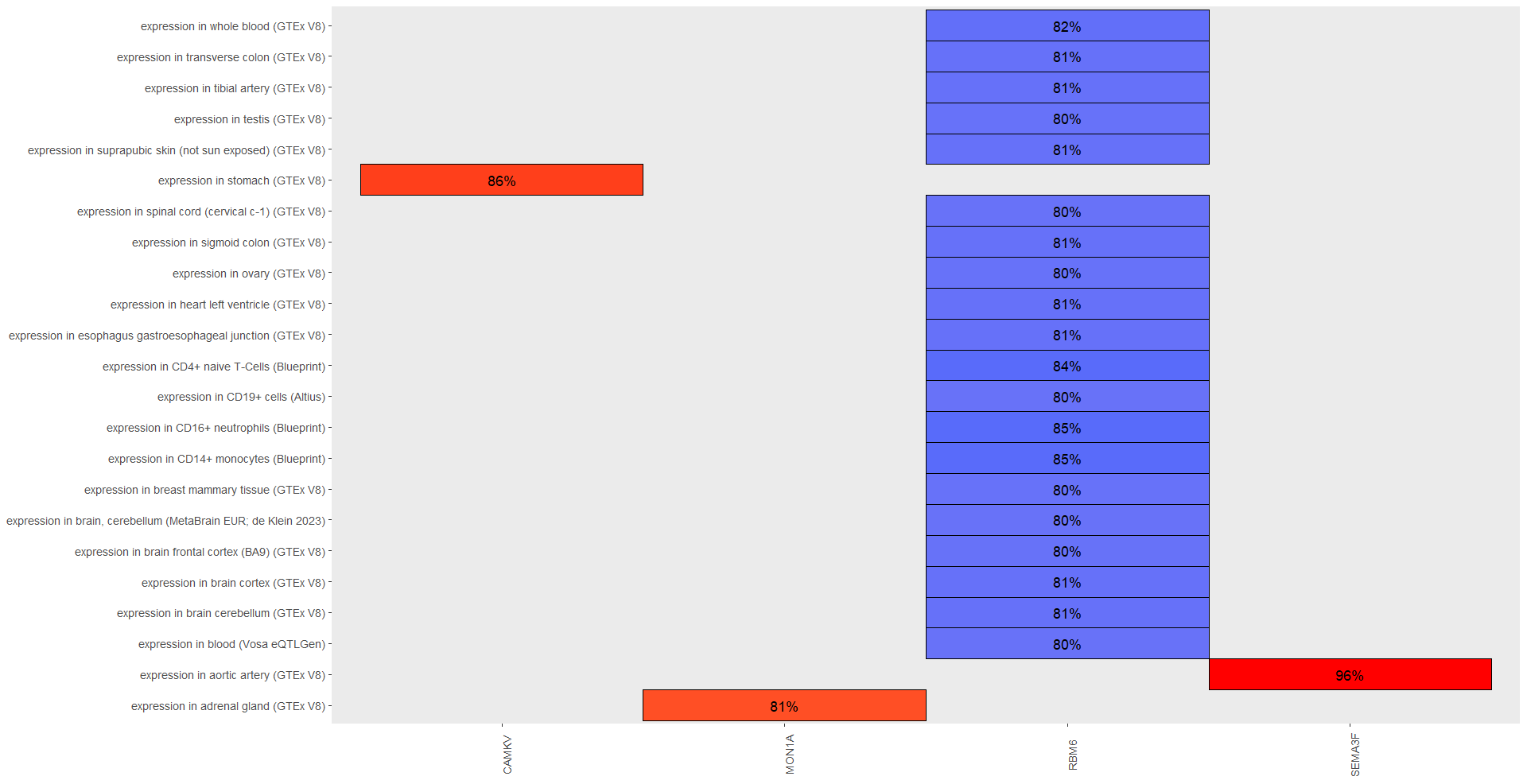

**A**

**B**

*A: Each dot represents a SNP with the x-axis showing the chromosomal location of the SNP and the y-axis representing the strength of association by the -log_10_(P-value)*. The SNPs coloured in red are contained within the 95% credible sets. *The variant circled is the SEMA3F missense variant (Leu504Met) which is in the credible set of the signal (rs1046956:T:A, EAF = 0.72, OR = 0.96, P = 1.4 x 10^-13^). B: Colocalisation plot highlighting significant colocalisations (PP>0.8) between RNA expression of genes within the locus (x-axis) and chronic pain susceptibility. Red indicates that higher RNA expression is associated with increased risk of chronic pain, whilst blue indicates that higher RNA expression is associated with decreased risk of chronic pain.*

**Supplementary Figure 6: Regional plots of *PER3* association with pain susceptibility (strong opioid use for >90 days vs healthy) in combined analyses (Panel A), female-only analysis (Panel B)**, **and male-only analysis (Panel C)**

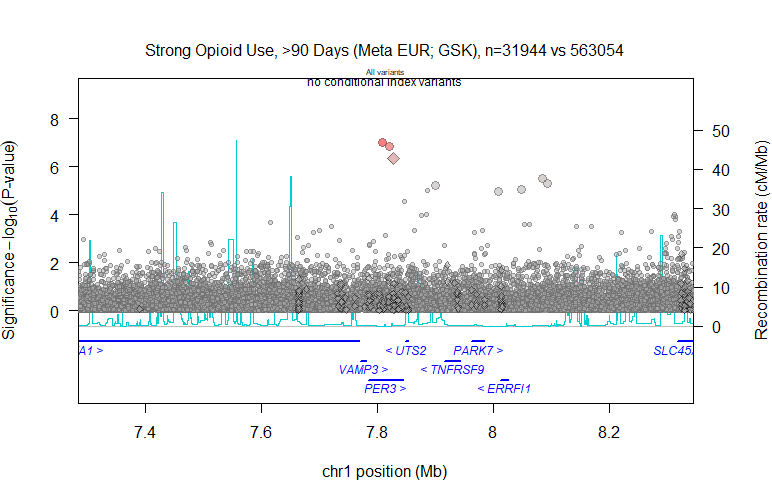

**A**

**B**

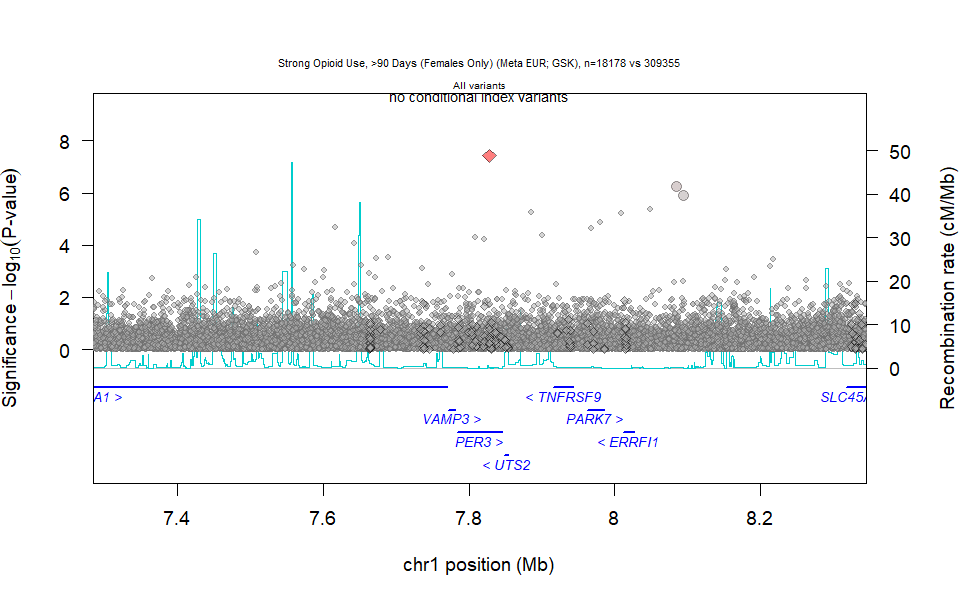

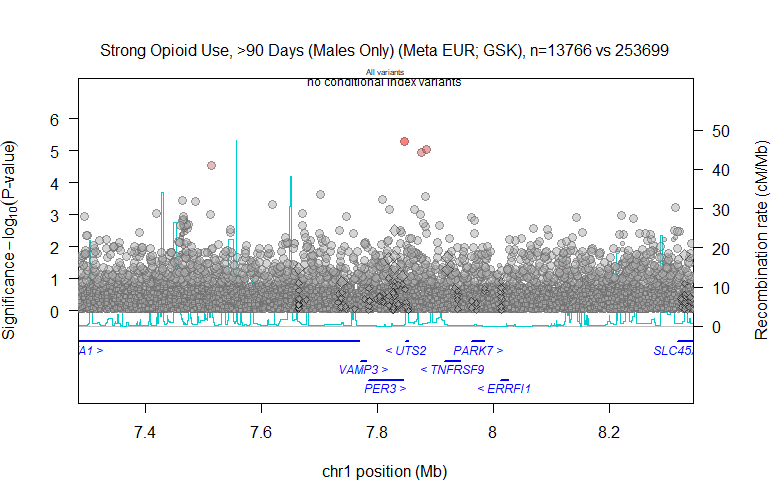

**C**

*Each dot represents a SNP with the x-axis showing the chromosomal location of the SNP and the y-axis representing the strength of association by the -log_10_(P-value)*. The SNPs coloured in red are contained within the 95% credible sets. A diamond indicates a missense variant.

**Supplementary Figure 7: Regional plots of *OPRM1* association with pain susceptibility (time on analgesics Q4 vs healthy) in combined analyses (Panel A), female-only analysis (Panel B),** **and male-only analysis (Panel C) and pain duration (time on analgesics Q4 vs Q1-Q3) in combined analyses (Panel D), female-only analysis (Panel E), and male-only analysis (Panel F).**

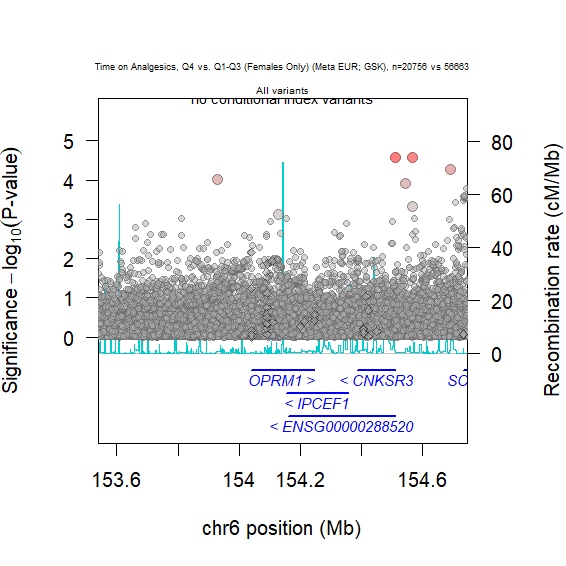

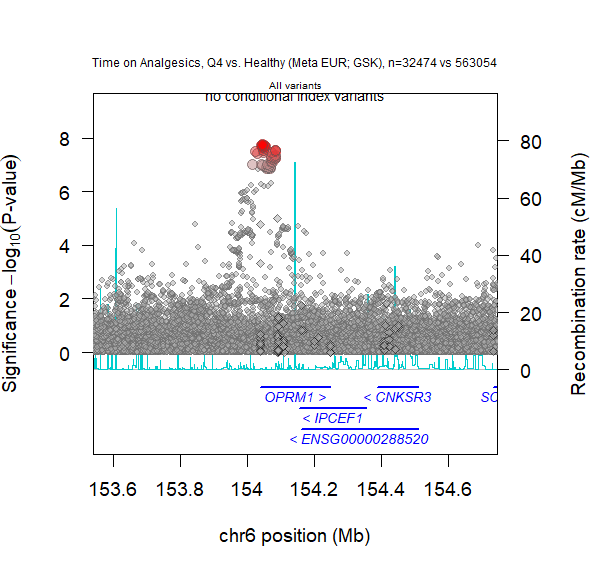

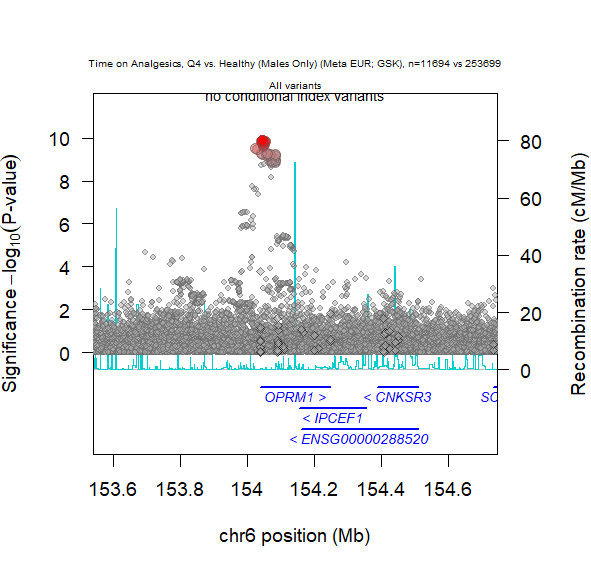

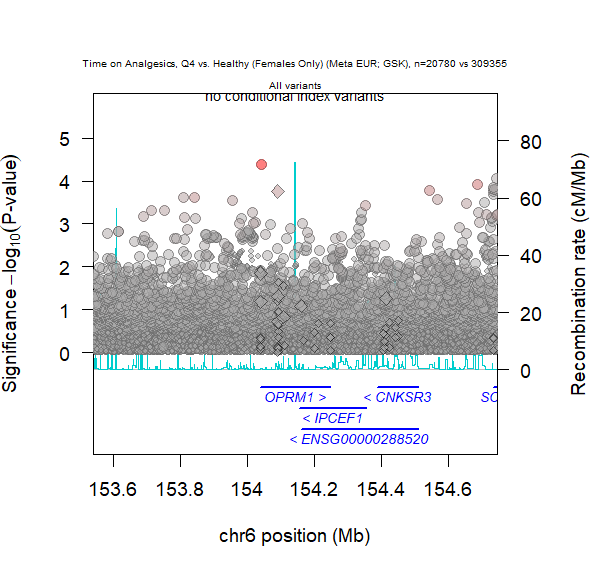

**A**

**B**
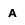

**C**
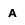
**C**

**E**
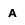
**E**

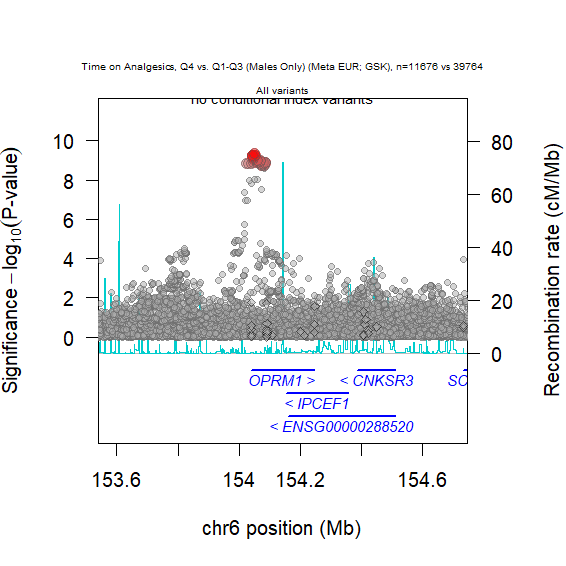

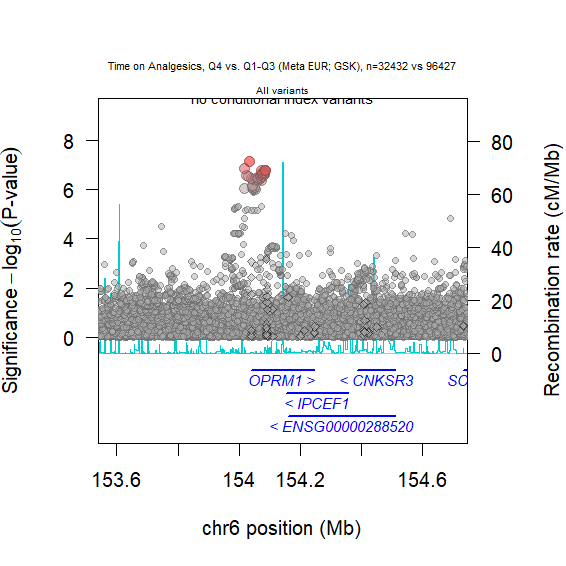

**F**
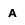
**C**

**D**

*Each dot represents a SNP with the x-axis showing the chromosomal location of the SNP and the y-axis representing the strength of association by the -log_10_(P-value)*. The SNPs coloured in red are contained within the 95% credible sets.
